# The mutagenic forces shaping the genomic landscape of lung cancer in never smokers

**DOI:** 10.1101/2024.05.15.24307318

**Authors:** Marcos Díaz-Gay, Tongwu Zhang, Phuc H. Hoang, Azhar Khandekar, Wei Zhao, Christopher D. Steele, Burçak Otlu, Shuvro P. Nandi, Raviteja Vangara, Erik N. Bergstrom, Mariya Kazachkova, Oriol Pich, Charles Swanton, Chao Agnes Hsiung, I-Shou Chang, Maria Pik Wong, Kin Chung Leung, Jian Sang, John McElderry, Lixing Yang, Martin A Nowak, Jianxin Shi, Nathaniel Rothman, David C. Wedge, Robert Homer, Soo-Ryum Yang, Qing Lan, Bin Zhu, Stephen J. Chanock, Ludmil B. Alexandrov, Maria Teresa Landi

## Abstract

Lung cancer in never smokers (LCINS) accounts for up to 25% of all lung cancers and has been associated with exposure to secondhand tobacco smoke and air pollution in observational studies. Here, we evaluate the mutagenic exposures in LCINS by examining deep whole-genome sequencing data from a large international cohort of 871 treatment-naïve LCINS recruited from 28 geographical locations within the Sherlock-*Lung* study. *KRAS* mutations were 3.8-fold more common in adenocarcinomas of never smokers from North America and Europe, while a 1.6-fold higher prevalence of *EGFR* and *TP53* mutations was observed in adenocarcinomas from East Asia. Signature SBS40a, with unknown cause, was found in most samples and accounted for the largest proportion of single base substitutions in adenocarcinomas, being enriched in *EGFR*-mutated cases. Conversely, the aristolochic acid signature SBS22a was almost exclusively observed in patients from Taipei. Even though LCINS exposed to secondhand smoke had an 8.3% higher mutational burden and 5.4% shorter telomeres, passive smoking was not associated with driver mutations in cancer driver genes or the activities of individual mutational signatures. In contrast, patients from regions with high levels of air pollution were more likely to have *TP53* mutations while exhibiting shorter telomeres and an increase in most types of somatic mutations, including a 3.9-fold elevation of signature SBS4 (q-value=3.1 × 10^−5^), previously linked mainly to tobacco smoking, and a 76% increase of clock-like signature SBS5 (q-value=5.0 × 10^−5^). A positive dose-response effect was observed with air pollution levels, which correlated with both a decrease in telomere length and an elevation in somatic mutations, notably attributed to signatures SBS4 and SBS5. Our results elucidate the diversity of mutational processes shaping the genomic landscape of lung cancer in never smokers.

## INTRODUCTION

While cancers of the lungs are commonly associated with tobacco smoking^1^, prior studies have indicated that between 10 and 25% of the 2.2 million lung cancer cases worldwide are found within individuals that have never smoked tobacco cigarettes^2,3^. The prevalence of lung cancer in never smokers (LCINS) varies by several notable factors, namely enrichment in females^4^, Asian populations^5^, and individuals with a family history of lung cancer^6^. The occurrence of LCINS can also differ by geographic regions, with higher rates reported in East Asia^7,8^ and Eastern Europe^9^ when compared to countries in North America and Western Europe^10^. Epidemiological studies have also identified environmental exposures that can increase LCINS risk, including exposure to secondhand smoke^11^ and air pollution^12,13^.

Elucidating the mutational signatures operative within a cancer genome allows the understanding of mutational processes implicated in cancer development^14^. Prior analyses have revealed more than 100 characteristic signatures across the spectrum of human neoplasia, with putative etiology assigned to approximately one third of known signatures^15^. However, there has been no comprehensive examination of the mutational signatures operative in LCINS as prior lung cancer whole-genome sequencing studies have focused almost exclusively on tobacco smokers from European descent^16,17^ and they have included only a relative small number of LCINS, mostly from East Asian descent^18^. To the best of our knowledge, our prior whole-genome sequencing study encompassing 232 lung cancers from never-smokers^19^, which was focused on evolutionary classification and its potential for personalized treatment, has been the largest examination of the mutational landscape in LCINS.

To understand the mutational processes shaping the lung cancer genomes of never smokers, here, we generated and analyzed deep whole-genome sequencing data from 871 treatment-naïve LCINS, including lung cancer histologies both commonly and rarely found in never-smokers. Our large international cohort of LCINS allowed a comprehensive evaluation of the mutagenic role of secondhand smoke, as well as the examination and comparison of mutational signatures between biological sexes, ancestries, and 28 geographic regions with different levels of air pollution.

## RESULTS

### The Sherlock-*Lung* never-smoking lung cancer cohort

As part of the Sherlock-*Lung* study^20^, a total of 871 treatment-naïve never-smoking lung cancer cases were whole-genome sequenced with a mean coverage of 88x and 35x for tumor and matched germline samples, respectively. Patients were recruited from 28 different locations across four continents (**Fig. 1*a***) and were predominantly female (*n=*688; 79.0%). Patients from North America and Europe (NA/EU; *n*=541; 62.1%) mostly clustered with the EUR super-sample from the 1,000 Genomes Project^21^ (472/541; 87.2%), and patients from East Asia (AS; *n*=309; 35.5%) clustered exclusively with the EAS super-sample (**Fig. 1*b***). Patients of European descent were almost exclusively recruited from Europe, Russia, USA, and Canada, while patients from Asian descent were recruited mainly in Hong Kong, Taipei, Korea, and Canada (**Supplementary Table 1**). Most patients were diagnosed with adenocarcinomas (*n*=737; 84.6%) or carcinoid tumors (*n*=61; 7.0%). In addition, we report the genomic landscapes of 31 squamous cell carcinomas and additional rarer histologies, including 13 adenosquamous carcinomas, 5 large cell carcinomas and 24 tumors with uncertain histological subtype (**Supplementary Table 1**). Information on passive smoking was collected for 458 patients, with 250 exposed and 208 not exposed to secondhand tobacco smoke. No subjects included in the study reported occupational exposure to mutagenic agents.

**Fig. 1.**
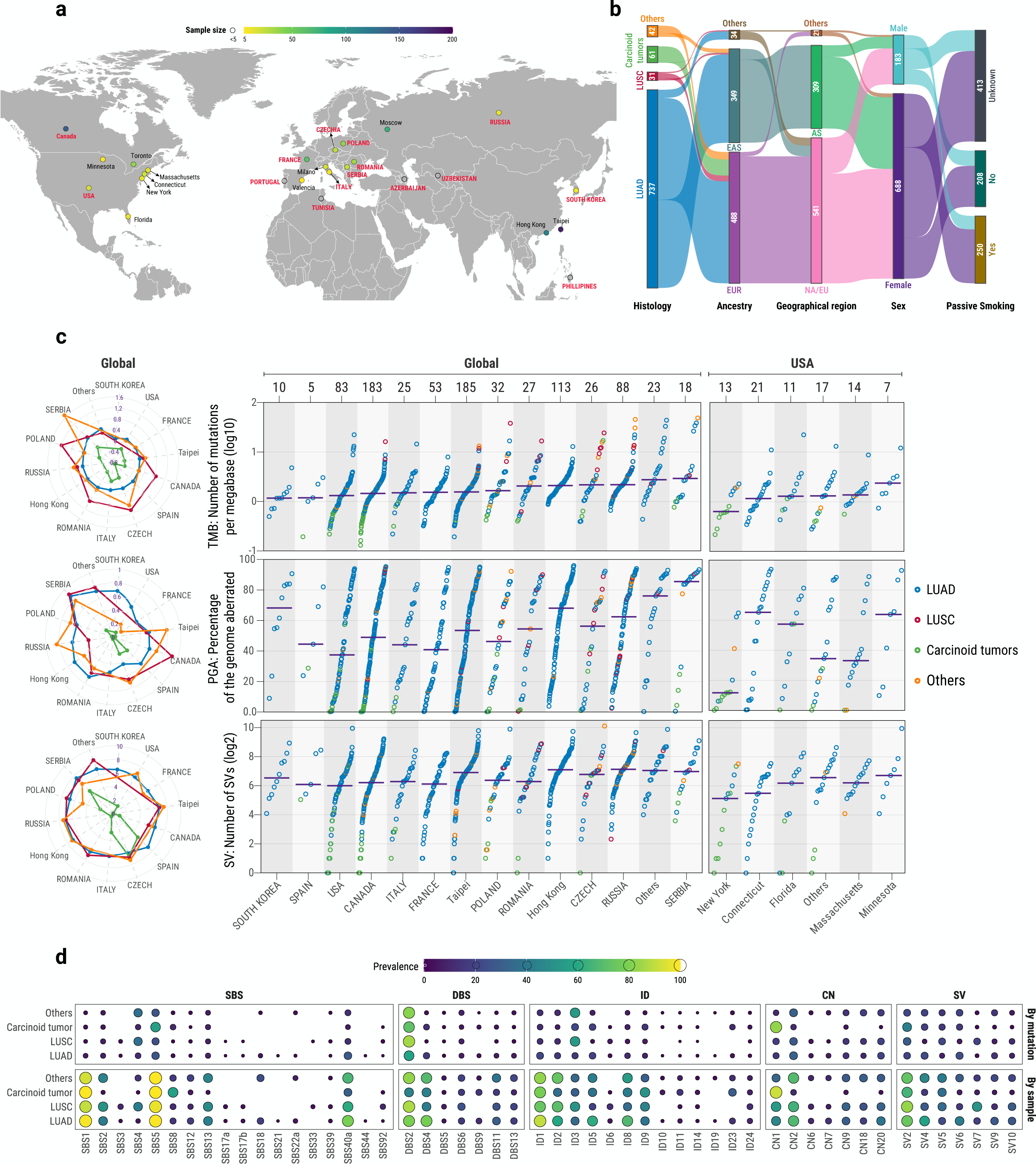
Overview of the Sherlock-*Lung* cohort of lung cancers in never smokers. **a**, Geographical distribution of the 871 patients across four continents and 28 geographic locations. **b**, Clinical characterization based on histology, genetic ancestry, geographical region, biological sex, and passive smoking status. **c**, Prevalence of mutations, percentage of genome altered, and structural variants across geographic locations and stratified based on histology. Left panel, dots represent median values for the three genomic alterations individually per country and histology. Right panel, dots represent individual tumors, colors different histology types, and horizontal purple lines median values across all histologies. **d**, Landscape of mutational signatures across histologies and somatic variant classes, including single base substitutions (SBS), doublet base substitutions (DBS), small insertions and deletions (ID), copy number alterations (CN), and structural variants (SV). The size and color of the dots represent the percentage of mutations contributed by the signature in all samples sharing the same histology (top panel) or the percentage of samples of a histological type where a particular signature is active (bottom panel). AS: East Asian geographical regions, EAS: East Asian genetic ancestry super-sample, EUR: European genetic ancestry super-sample, LUAD: lung adenocarcinomas, LUSC: lung squamous cell carcinomas, NA/EU: North America and Europe geographical regions.

### Mutational patterns of lung cancer in never smokers

The median tumor mutational burden (TMB) was 5,068 for single base substitutions (SBSs; range 393-151,385), 279 for small insertions and deletions (IDs; range 14-10,474), and 105 for structural variants (SV; range 1-1,103 **Fig. 1*c***), with differences across histologies and geographical locations. Similarly, the median percentage of genome aberration was 55.3%, with clear variances across histologies and geographical areas (**Fig. 1*c***). To identify the mutational processes operative in LCINS, we performed *de novo* extraction^22^ of mutational signatures for SBS, ID, doublet base substitutions (DBS), copy number (CN) alterations, and structural variants (SV; **Supplementary Tables 2-6**). Eleven *de novo* SBS signatures were extracted and further decomposed into 18 previously identified signatures from the Catalogue of Somatic Mutations in Cancer (COSMICv3.4) reference signature database^23^ (**Supplementary Fig. 1**; **Supplementary Table 7**). The unprecedented size of the cohort allowed us to refine the prevalence and intensity of the signatures involved in LCINS, with several signatures identified for the first time in LCINS, including signature SBS40a, recently extracted in clear cell renal cell carcinomas but shown to be active in multiple cancer types^15,24^. Other SBS signatures not previously seen in LCINS included unknown etiology signatures SBS12, SBS33, and SBS39 as well as the aristolochic acid-related signature SBS22a and tobacco smoking-associated signatures SBS4 and SBS92 (**Fig. 1*d***). Additionally, the mismatch repair deficiency-associated signatures SBS21 and SBS44 were detected in one lung adenocarcinoma case, which had 10,474 indels and was confirmed by MMRDetect^25^ as microsatellite unstable cancer. Analysis of indels and doublet base substitutions revealed a new compendium of mutational signatures operative in LCINS (**Supplementary Figs. 2-3**; **Supplementary Tables 8-9**). Specifically, a novel ID signature was identified (termed, ID24), characterized by short microhomology deletions, whereas several ID and DBS signatures were identified for the first time in LCINS, including ID6, ID10, ID11, ID14, ID19, ID23, DBS5, DBS6, and DBS13. *De novo* extraction of signatures of large mutational events revealed four CN (**Supplementary Fig. 4**) and four SV (**Supplementary Fig. 5**) signatures, which were subsequently decomposed into the recently described COSMICv3.4 reference signatures^26-29^ (**Supplementary Tables 10-11**).

### Mutational landscape of lung adenocarcinoma in never smokers

The genomic landscape of the 737 lung adenocarcinomas from never smokers showed a remarkable heterogeneity for both SBS and CN alterations (**Fig. 2*a***). Approximately 4.9% of adenocarcinomas (36/737) harbored tobacco-associated signature SBS4, while the most prevalent signature was SBS40a, of unknown etiology, which contributed 28.2% of all substitutions. Interestingly, a cluster of 25 hypermutated samples (>25,000 mutations) with high proportions of SBS4 and APOBEC-associated signatures SBS2 and SBS13 was identified (**Fig. 2*a-b***). Additionally, a smaller subset of 11 cases harboring signatures SBS3 and ID6, both linked to homologous recombination deficiency (HRD), was also identified. The presence of HRD was further evaluated using CHORD^30^ and HRDetect^31^, which jointly predicted six of the cases as HRD, with two additional cases predicted as HRD exclusively by CHORD (**Supplementary Fig. 6**).

**Fig. 2.**
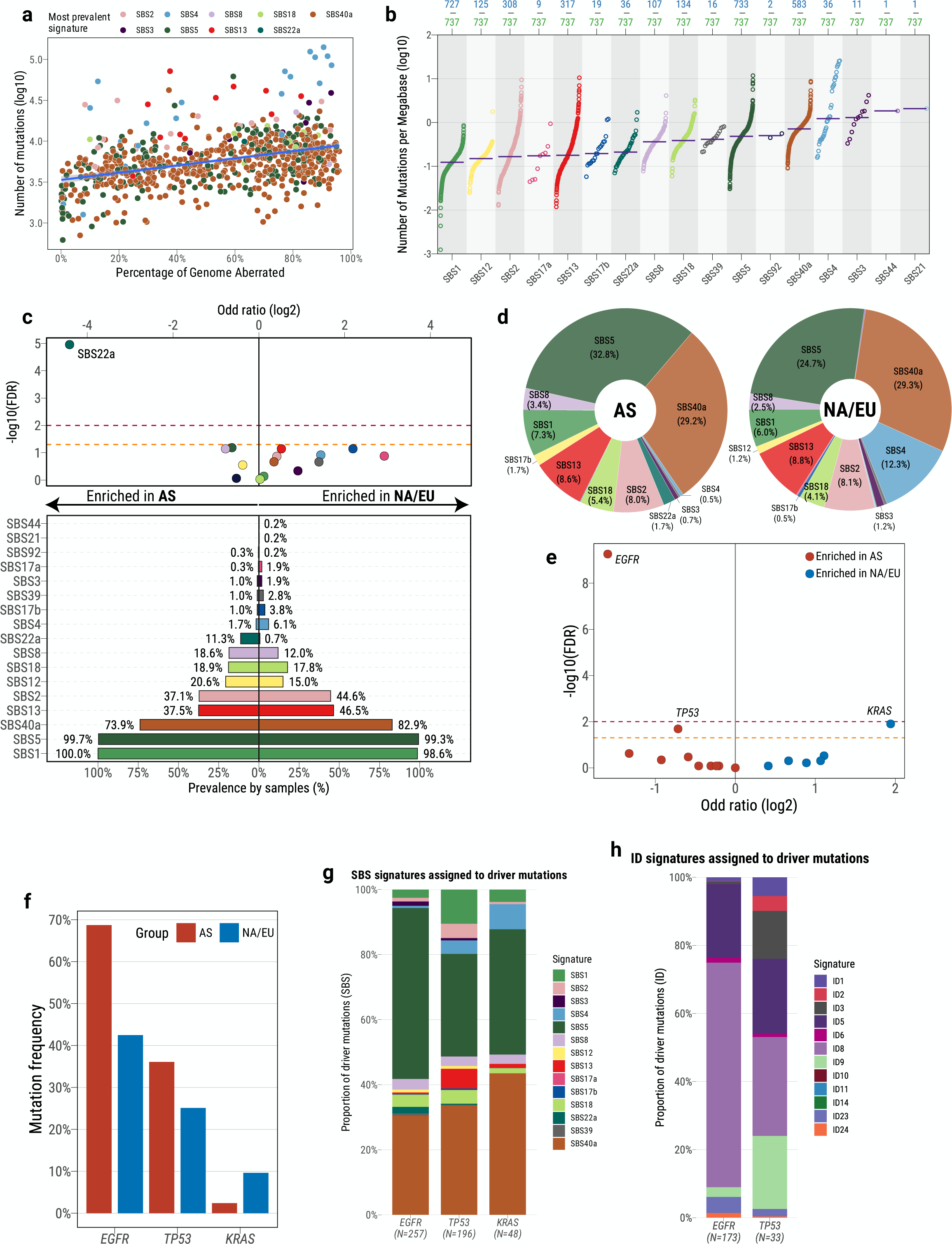
Repertoire of mutational signatures and driver mutations in LCINS adenocarcinomas. **a**, Distribution of the most prevalent signatures per sample according to the number of single base substitutions (SBS) and percentage of genome aberrated. **b**, Activity of SBS mutational signatures across samples, representing the total number of mutations attributed to each signature in a given sample. Dots represent individual samples and purple horizontal bars median values. The numbers on the bottom indicate the total number of samples where a particular signature was found active (blue) and the total number of LCINS adenocarcinoma samples (green). **c**, Regional differences across signatures. Volcano plot indicating enrichment of SBS signatures in patients from East Asian (AS) and North American/European regions (NA/EU) in LCINS adenocarcinomas (top panel) and bar plot indicating prevalence by geographical region (bottom panel). Horizontal lines marking statistically significant thresholds were included at 0.05 (dashed orange line) and 0.01 FDR levels (dashed red line). **d**, Total number of mutations assigned to specific SBS signatures for patients from East Asia or North America and Europe. **e**, Volcano plot indicating enrichment of mutations in driver genes affecting specific LCINS adenocarcinomas. Blue-colored genes were enriched in patients from North America and Europe, whereas red-colored genes were enriched in patients from East Asia. **f**, Frequency of LCINS adenocarcinoma cases harboring driver mutations in the driver genes significantly differently mutated between geographical regions (*EGFR*, *TP53*, and *KRAS*) **g**, Proportion of driver mutations affecting *EGFR*, *TP53*, and *KRAS* probabilistically assigned to each of the SBS mutational signatures identified in the LCINS adenocarcinoma cohort. The numbers indicate the total number of driver single base substitutions found in each of the genes.

Comparisons between patients from different regions revealed differences in somatic mutagenesis (**Fig. 2*c-d***). Notably, signature SBS22a (previously known as SBS22, and recently renamed^24^) was enriched in patients from East Asia (**Fig. 2*c***). This enrichment was also observed after removing the patients from Canada with EAS ancestry (**Supplementary Fig. 7**; **Methods**), which encompassed most of the patients with EAS ancestry not residing in Asia (35/40, 87.5%; **Fig. 1*b***). Further, SBS22a was found almost exclusively in patients from Taipei (32/36 SBS22a-positive cases, 88.9%). Although SBS22a has been associated with aristolochic acid exposure in liver, bladder, and kidney cancers^32,33^, this is the first evidence of aristolochic acid causing mutations in lung cancer. Moreover, SBS22a also showed a significant co-occurrence with SBS12 in patients from East Asia (OR=4.55; p-value=1.2 × 10^−4^). SBS12 is commonly found in gastrointestinal cancers, especially cancers of the livers^15,34^ and kidneys^24^, with a consistent enrichment of the signature in patients from East Asia^24,34^.

Several signatures of indels and large genomics alterations differed across regions (**Extended Data Fig. 1a**), with ID3, CN20, SV2, SV4, SV6, and SV9 being more prevalent in patients from East Asia, whereas three additional SV signatures, all linked primarily with non-clustered alterations, SV5, SV7, and SV10, were enriched in North American and European patients. Interestingly, CN20 was previously found elevated in Black TCGA patients but not in Asian patients when compared to White patients^26^. Additionally, signatures SBS4 and ID3 showed a difference between males and females across the spectrum of variant classes, with a notably higher prevalence in males, whereas ID9 was enriched in females (**Extended Data Fig. 1b**).

As in prior LCINS studies^10^, *EGFR* and *TP53* harbored the most driver mutations, with 52.2% and 30.5% mutated in adenocarcinomas, respectively, whereas *KRAS* was only mutated in 6.5% of samples. *EGFR* and *TP53* mutations were enriched in patients from East Asia, as previously reported^35^, and *KRAS* mutations were elevated in North American and European patients (**Fig. 2e-f** and **Extended Data Fig. 1c**-e). No differences were found for driver mutations between males and females for adenocarcinomas (**Extended Data Fig. 1f**). Interestingly, for *EGFR*-mutated tumors, a significant increase in TMB was observed for those *TP53*-wild-type (q-value=1.2 × 10^−4^), whereas a decrease was found for *TP53*-mutated cases (q-value=0.15; **Extended Data Fig. 1g**). In addition, while analyzing *EGFR*-mutated cases specifically, we identified a high enrichment in the prevalence of signature SBS40a, along with other signatures of multiple variant types (**Extended Data Fig. 1h**).

Most driver mutations in *EGFR*, *TP53*, and *KRAS* were probabilistically assigned to signatures SBS5 and SBS40a (**Fig. 2*g***), with the proportion of driver mutations significantly higher than expected according to their prevalence in the overall adenocarcinoma cohort (OR=2.55; p-value=9.8 × 10^−22^; **Fig. 2*d***). Prior analyses have shown that SBS4 generates the majority of driver mutations in *KRAS* in tobacco smokers^36^. In our samples, SBS4 contributed only ∼10% of KRAS mutations. Most driver indels, especially in *EGFR* (mostly exon 19 deletions; 150/173, 86.7%), were probabilistically assigned to signature ID8 (**Fig. 2h**), which has been previously related to radiation exposure^37^. Despite contributing a significant number of mutations in adenocarcinomas (**Fig. 2*d***), APOBEC-associated signatures SBS2 and SBS13 generate a significantly lower number of driver mutations in *EGFR*, *TP53*, and *KRAS* (OR=0.28; p-value=2.8 × 10^−14^) in comparison to their prevalence in the overall adenocarcinoma cohort, with most APOBEC-associated mutations being *TP53* point mutations in a small number of samples (**Fig. 2*g***).

### Mutational landscape of lung carcinoids in never smokers

Carcinoid tumors were the second most common histology, with all samples originating from North American and European patients except one case from Taipei (**Supplementary Table 1**), and only one case harboring signature SBS4. Carcinoids presented a lower number of single base substitutions (p-value=3.3× 10^−29^), copy number (p-value=2.6× 10^−32^), and structural variants (p-value=4.2× 10^−26^) compared to adenocarcinomas, as well as longer telomeres (p-value=2.0× 10^−6^; **Extended Data Fig. 2**). SBS5 accounted for 55.7% of all mutations in carcinoids in contrast to 27.0% in the adenocarcinomas (p-value<2.2× 10^−16^; **Fig. 1*d*** and **Extended Data Fig. 3a**). In comparison to adenocarcinomas, carcinoids showed 3.39-fold lower TMB (**Extended Data Fig. 2a**), with depletion of signatures SBS2, SBS5, SBS12, and SBS13 (**Extended Data Fig. 4a**). Nevertheless, we observed an enrichment in signature SBS8, previously linked to nucleotide excision repair^38^ and late replicating regions^39^. Interestingly, SBS8 was a relatively minor signature in adenocarcinomas (**Fig. 1*d***) but present in the majority of carcinoids (35/61), where it contributed 11.0% of all mutations (**Extended Data Fig. 3a**). As lung carcinoids are thought to originate from neuroendocrine cells producing hormones and hormone-like substances^40^, the presence of SBS8 is perhaps not surprising as this mutational signature is commonly observed in hormone-dependent cancers such as breast^41^ and prostate^15^ adenocarcinomas.

Additionally, three ID signatures and the diploid CN1 signature were found enriched in carcinoids, whereas several different signatures were enriched in adenocarcinomas, including two DBS, four ID, and most CN and SV signatures (**Extended Data Fig. 3b** and 4a), as expected due to the lower number of large genomic aberrations of this histological subtype of LCINS (**Extended Data Fig. 2a**). No *EGFR* or *KRAS* driver mutations were found in carcinoid histology, whereas an enrichment was found for driver mutations in *ARID1A* (**Extended Data Fig. 3c** and 4b***-d***).

### Mutational landscape of lung squamous cell carcinomas in never smokers

Squamous cell carcinomas accounted for 3.6% of LCINS (31/871), with 13 samples harboring tobacco-associated signature SBS4. In comparison with adenocarcinomas, squamous cell carcinomas showed an elevated burden of SBS, DBS, and ID, in contrast to a similar landscape of large genomic alterations and telomere lengths (**Extended Data Fig. 2**). Several signatures showed higher prevalence compared to adenocarcinomas, including SBS3, SBS4, SBS92, and SV7 (**Extended Data Fig. 5a**-c and **6*a***). *TP53* was the most prevalent driver mutation (58.1% of cases; **Extended Data Fig. 5d**) and was found enriched in LCINS squamous cell carcinomas compared to adenocarcinomas, along with mutations in *LRP1B*, *PIK3CA*, and *PTEN* (**Extended Data Fig. 6b**-g). In contrast, *EGFR* mutations were rarely observed in squamous cell carcinomas, as previously reported^42^ (**Extended Data Fig. 6d**).

### SBS4 in lung tumors from never smokers

Tobacco-associated signature SBS4 was found active in 56 LCINS tumors (6.4%), and was the predominant signature in 39 of them (69.6%) and in many hypermutated cases (5.51-fold higher median SBS burden in SBS4+ cases; **Extended Data Fig. 7a**-b). Similarly, signature SBS92, recently linked to tobacco smoking in bladder^22,43^ and other cancer types^44^, was also active in four SBS4+ tumors, and found significantly enriched in SBS4+ cases (p-value=2.1 × 10^−4^; **Extended Data Fig. 7b**). Tumors presenting active SBS4 also showed a significant enrichment of previous ID and DBS signatures linked to tobacco smoking, namely ID3 (p-value=1.1 × 10^−22^) and DBS2 (p-value=5.0 × 10^−7^; **Extended Data Fig. 7c**-d). Additionally, SBS4 in LCINS exhibited the same genome topography characteristics as SBS4 in smokers^45^, including enrichment in late replicating regions, periodicity in the vicinity of nucleosomes, and strong genic and transcriptional strand asymmetries (**Extended Data Fig. 8**).

In principle, detecting SBS4 in never-smokers can be due to misannotation of tobacco smoking status. The observed SBS4 prevalence increase in males and lung squamous cell carcinomas indicates that some samples might have been incorrectly annotated as never smokers. Nevertheless, the 56 LCINS tumors with SBS4 exhibited statistically different driver mutations when compared to previously generated lung cancers from tobacco smokers^36^ (**Extended Data Fig. 7e**). Specifically, the LCINS samples had fewer mutations in *KRAS* (OR=0.41; p-value=0.021) and an enrichment of *EGFR* p.L858R hotspot mutations (OR=29.35; p-value=4.1 × 10^−4^). These differences indicate that, in addition to possible misannotation, other exogenous processes may be generating SBS4 in these never smokers. As prior work has shown that SBS4 can be found in lifelong non-smokers exposed to different environmental mutagens, including occupational history of coal tar work^46^ and indoor air pollution^47^, we evaluated whether SBS4 in this cohort, as well as mutations in other samples, can be generated by either secondhand exposure to tobacco smoking or by high levels of air pollution.

### Passive smoking has low mutagenicity in LCINS

To understand the mutational processes activated by secondhand tobacco smoking, we compared the genomic landscapes of 250 LCINS from passive smokers to 208 LCINS from individuals who were not exposed to secondhand smoke. We observed an increase of SBS mutations in passive smokers (**Fig. 3*a***), as well as a decrease in tumor/normal telomere length ratio (**Fig. 3*b***). Although not significant, the directionality of these associations with passive smoking was consistent after adjustment for other covariates, encompassing age, sex, genetic ancestry, histology, and tumor purity – 8.3% increase and 5.4% decrease in the magnitude of regression coefficients after covariate correction, respectively (**Fig. 3*c-d***). Almost identical results were observed when we restricted the analyses to the lung adenocarcinomas from never smokers (**Supplementary Fig. 8**), and when the principal components from the ancestry analysis were used instead of the ancestry labels for adjustment (**Supplementary Table 12**; **Methods**). However, this increase in mutation burden was not specifically associated with any mutational signature (**Fig. 3*e***) or mutation type (**Fig. 3*f***). In addition, no significant differences were found for mutations in any cancer driver genes (**Fig. 3*g***), nor for the burden of ID, DBS, SV, or CN segments (**Extended Data Fig. 9a**-e) or the presence of signatures derived from these mutation types (**Extended Data Fig. 9f**). Amongst the 250 cases identified as exposed to secondhand smoke, only three (1.2%) displayed signature SBS4 (OR=0.62; p-value=0.71), each with a contribution exceeding 20% and at least 500 mutations attributed to SBS4. Moreover, only two of the 281 driver mutations found in secondhand smoke-exposed samples were assigned to SBS4 with a probability above 50%, including a *TP53* missense mutation (p.Val157Phe) and a missense mutation (p.Val409Leu) in *NF1*.

**Fig. 3.**
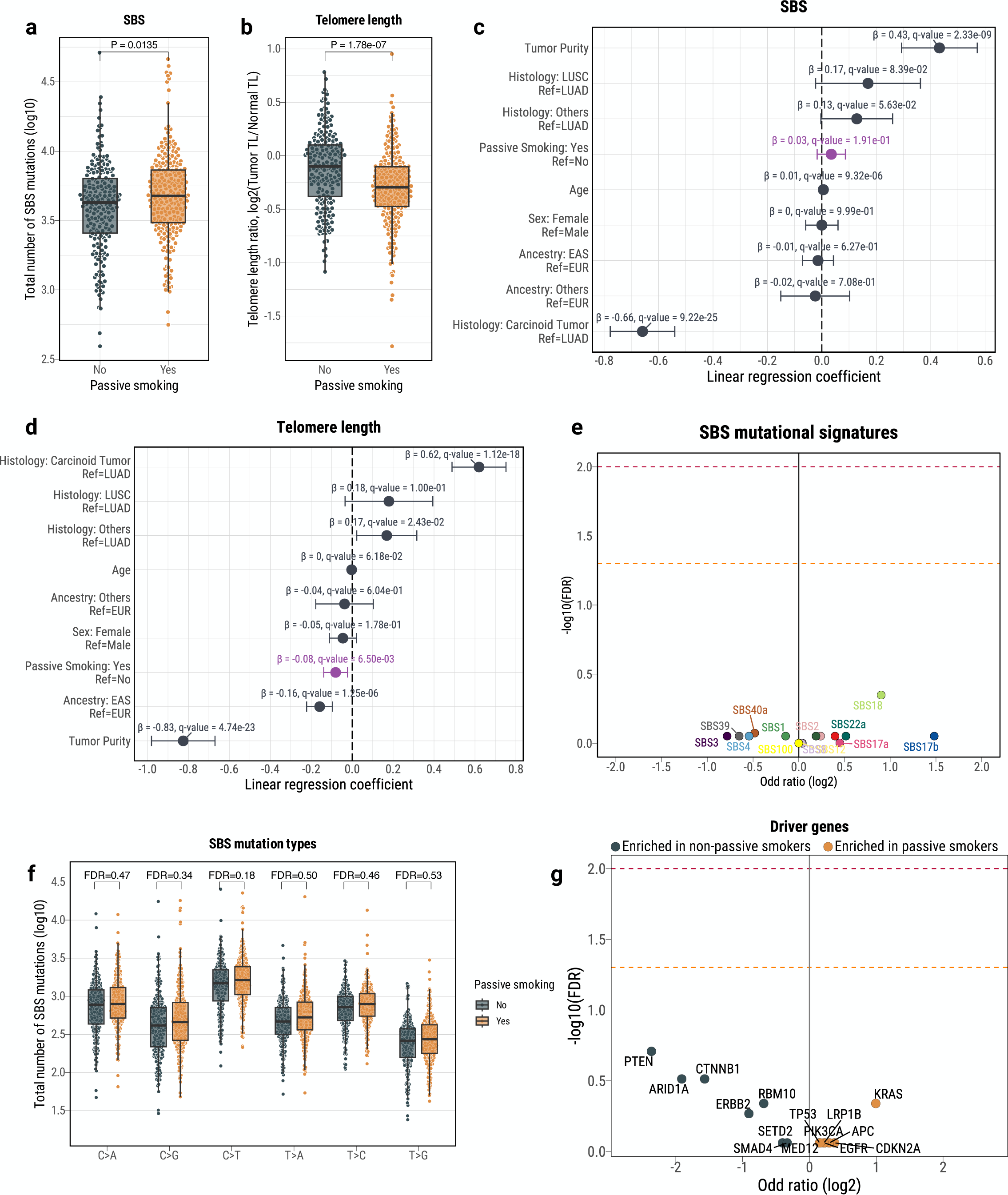
Passive smoking influence in the genomic landscape of LCINS. **a**-**d**, Differences in base substitution burden and tumor-to-normal telomere length ratio using univariate comparisons (**a** and **b**) as well as multivariable linear regressions considering clinical and epidemiological covariates (**c** and **d**), including age, sex, genetic ancestry, histology, and tumor purity. **e**, Volcano plot indicating enrichment of mutational signatures derived from SBS mutations in passive vs. non-passive smokers. Horizontal lines marking statistically significant thresholds were included at 0.05 (dashed orange line) and 0.01 FDR levels (dashed red line). **f**, Comparison of the mutations belonging to each of the six main SBS mutation subtypes in passive vs. non-passive smokers. **g**, Volcano plot indicating enrichment of mutations in driver genes affecting specific LCINS tumors.

To evaluate if we were unable to assign SBS4 to other secondhand smoke-exposed cases due to low levels of the signature, we synthetically injected SBS4 mutations using simulations at different levels into the mutational profiles of the 247 passive smoker cases that lacked SBS4 and assessed the number of samples where SBS4 was detected (**Supplementary Fig. 9**). We were able to detect SBS4 in around a quarter of simulated samples, if it contributed at least 5% of all mutations within a sample. Moreover, SBS4 was detectable in almost every sample when the signature accounted for more than 10% of mutations. These simulation results agree with the levels of SBS4 observed in the set of SBS4+ samples, where SBS4 is contributing above 10%. Thus, although SBS4 could have been missed in some secondhand smoke exposed LCINS, this signature would be likely contributing less than 5% of mutations in these cancers. Overall, our simulations demonstrate that it is unlikely that exposure to secondhand smoke accounts for SBS4, whereas the modest increase in overall SBS mutations suggests that passive smoking has low mutagenicity.

### Air pollution is associated with increased somatic mutations in LCINS

Given the recent evidence of the role of air pollution in lung carcinogenesis^12,48^, we assessed whether atmospheric air pollution, quantified by the environmental particulate matter measuring ≤2.5 μm (PM_2.5_), had an effect on the accumulation of somatic mutations in LCINS. Yearly PM_2.5_ estimates from 1998–2021 were obtained for the 853 LCINS cases with annotated country of residence using a hybrid model combining satellite-based measurements of aerosol optical depth with chemical transport modeling and ground-based observations (**Methods**)^49^. Considering the distribution of PM_2.5_ estimates across the cohort, a threshold of 20 μg/m^3^ was used to separate patients diagnosed in regions with high and low levels of pollution (**Supplementary Fig. 10**). LCINS originating from areas with high PM_2.5_ levels exhibited increased burdens of SBS (13.0%), DBS (8.3%), and ID mutations (7.6%; **Fig. 4*a***), along with telomere shortening (**Fig. 4*b***), even after accounting for age, sex, genetic ancestry, histology, and tumor purity (**Fig. 4*c-d***; **Supplementary Table 12**). No enrichments were identified for the numbers of SV or CN segments after corrections (**Extended Data Fig. 10a**-b). In order to determine whether higher levels of pollution lead to an increase in somatic mutagenesis (*i.e.*, a dose-response effect), we calculated individual estimates of PM_2.5_ per lung cancer patient (**Methods**). Consistent with a dose-response effect, we found statistically significant positive correlations of PM_2.5_ with the burden of SBS, DBS, and ID (**Fig. 4*e-g***), as well as a negative correlation with telomere length (**Fig. 4*h***), further corroborating the dose-response relationship of outdoor air pollution and somatic mutations in LCINS.

**Fig. 4.**
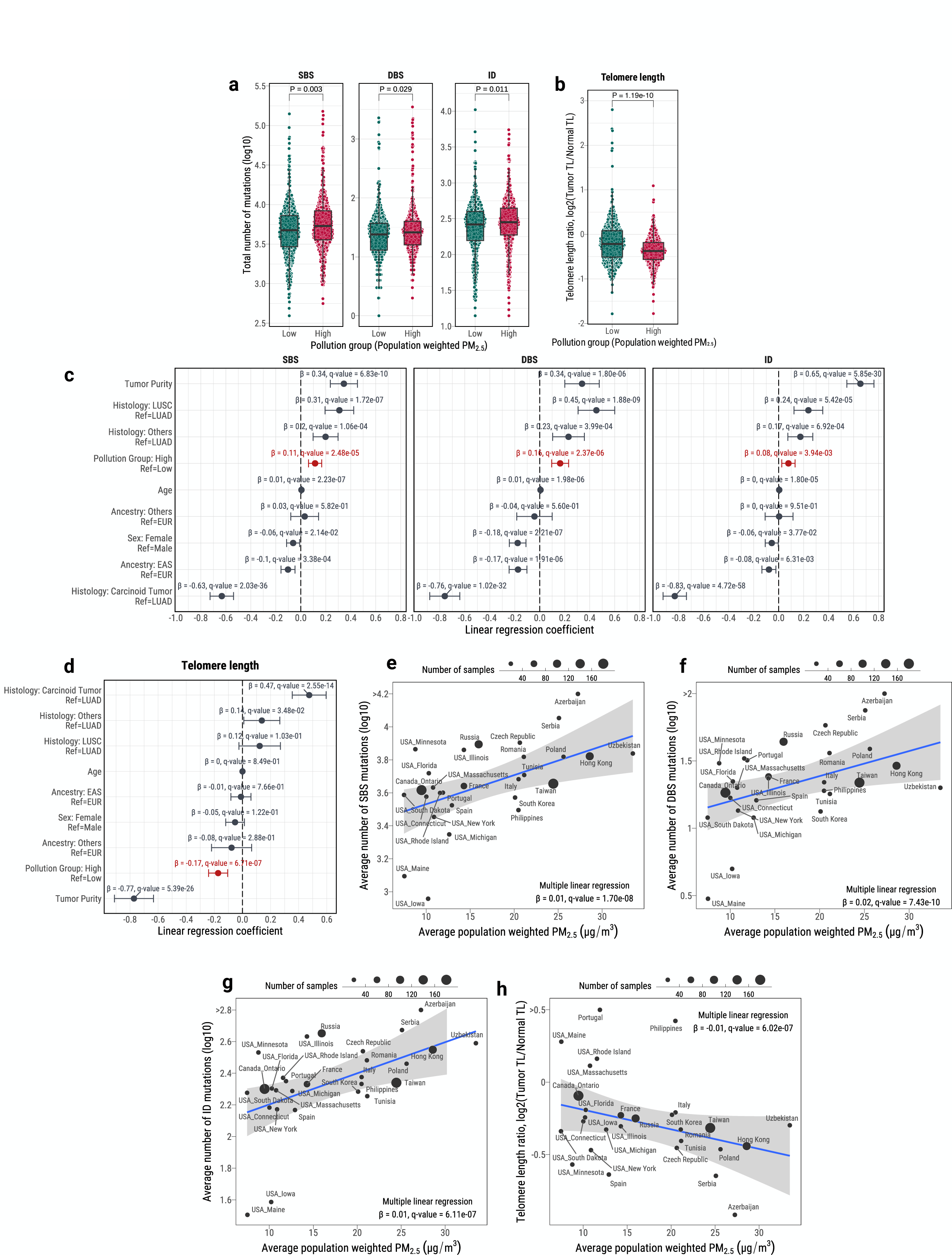
Mutagenic effects of PM_2.5_ exposure in LCINS. **a**, Quantification of tumor mutational burden according to different mutation types, including SBS, DBS, and ID, for patients living in geographical regions with high and low PM_2.5_ exposure levels (threshold defined at 20 μg/m^3^; only samples for which the country of origin was known (*n*=853) are included). **b**, Quantification of the ratio of telomere lengths for tumor and normal samples across high and low PM_2.5_ exposed cases. **c**-**d**, Forest plots corresponding to multivariable linear regressions considering high/low PM_2.5_ exposure group, age, sex, genetic ancestry, histology, and tumor sample purity as covariates and tumor mutational burden for the specific mutation type, SBS, DBS, ID (**c**), or telomere length ratio (**d**) as independent variables. **e**-**h**, Scatter plots showing significant correlations between individual sample estimates of PM_2.5_ exposure and tumor mutational burden for SBS (**e**), DBS (**f**), ID (**g**), and telomere length ratio (**h**).

Several mutational signature-specific associations with individual PM_2.5_ estimates were found after adjusting for covariates (**Fig. 5*a***). These included clock-like signature SBS5 (OR for 10 μg/m^3^of PM_2.5_=1.76; q-value=5.0 × 10^−5^) as well as signatures SBS4 (OR=3.86; q-value=3.1 × 10^−5^) and ID3 (OR=1.63; q-value=5.1 × 10^−4^; **Fig. 5*b***; **Supplementary Table 12**). Furthermore, dose-response effects were also observed for these signatures, with a significant positive correlation between the total numbers of mutations assigned to a given signature and the individual PM_2.5_ estimates (**Fig. 5*c-e***). Specifically, a unit (1μg/m^3^) increase of PM_2.5_ was associated with a 2.3% increase in SBS5-associated mutations, 12.0% in SBS4-associated mutations, and 6.0% in ID3-associated mutations, independent of other covariates. No associations were found with DBS, CN, or SV signatures (**Fig. 5*a***; **Extended Data Fig. 10c**). Patients in regions with high PM_2.5_ exposure were 1.6 times more likely to have *TP53* mutations and 2.5 times less likely to have *CTNNB1* mutations (**Fig. 5*f-h***). Overall, these results indicate that elevated air pollution levels are associated with an increase in both somatic mutations contributing to specific mutational signatures as well as an increased prevalence of *TP53* mutations.

**Fig. 5.**
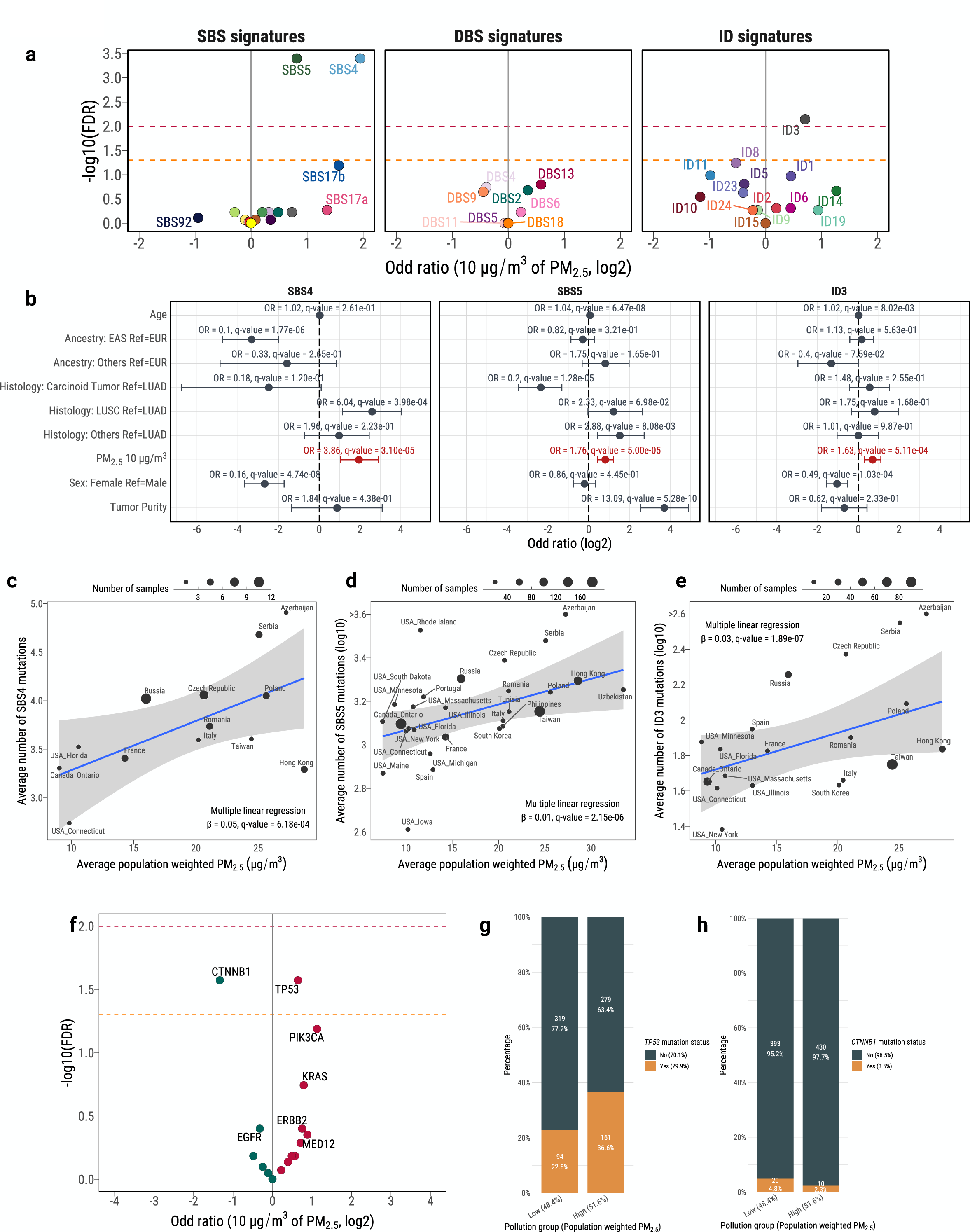
Associations between PM_2.5_ exposure and specific mutational signatures affecting LCINS tumors. **a**, Enrichment analysis of the presence of mutational signatures derived from SBS, DBS, and ID with PM_2.5_ exposure levels for all samples for which the country of origin was known (*n*=853). Horizontal lines marking statistically significant thresholds were included at 0.05 (dashed orange line) and 0.01 FDR levels (dashed red line). The odds ratios for the increase/decrease of 10 μg/m^3^ of PM_2.5_ estimates are shown. **b**, Detailed forest plots for the logistic regression models corresponding to signatures SBS4, SBS5, and ID3. **c-e**, Scatter plots assessing dose-response effect between individual sample estimates of PM_2.5_ exposure and mutations assigned to signatures SBS4 (**c**), SBS5 (**d**), and ID3 (**e**). **f**, Enrichment analysis of the presence of mutations in driver genes with PM_2.5_ exposure levels. **g**-**h**, Detail of the enrichment of *TP53* (**g**) and *CTNNB1* (**h**) driver mutations in high and low pollution regions, respectively.

## DISCUSSION

This study presents the most extensive mutational analyses of whole-genome sequenced lung cancers in never smokers. Our results reveal multiple mutational signatures and mutated cancer driver genes whose frequencies differ across lung cancer histologies, regions, and exposures. Particularly noteworthy is our finding of the aristolochic acid signature SBS22a in patients from Taipei, possibly shedding light on one of the previous unappreciated environmental factors contributing to the accumulation of mutations in LCINS from East Asia. Similarly, multiple signatures of indels and large genomics alterations, most with unknown etiology, were found to differ across regions, further highlighting the likely existence of additional population-specific exposures and other factors contributing to LCINS. Notably, SBS40a, with an unknown cause, and previously associated with kidney cancer^24^, accounted for the largest proportion of substitutions in adenocarcinomas, thus indicating the existence of a globally widespread and previously unappreciated mutagenic process, likely of endogenous origin.

Prior epidemiological research^11^ has established that passive smoking can be associated with a modest heightened risk for developing LCINS. In line with the observational research, our analysis of 458 patients with known exposure to passive smoking uncovered only minimal indications of elevated mutagenesis amongst individuals exposed to secondhand smoke. As in previous epidemiological studies, the data about second-hand tobacco smoking exposure was derived from questionnaires and clinical datasets, and varied across individuals, going from very detailed (during childhood, at work, or with a smoking spouse) to just binary yes/no exposure status. Thus, low exposure levels or very remote exposure (during childhood) may contribute to our findings of low mutagenicity. Nevertheless, our results are also consistent with a recent study in 291 Japanese LCINS patients with extensive information on passive smoking^50^. Additional non-mutagenic carcinogenesis could be contributing to the observed rise in lung cancer in those exposed to secondhand smoke.

Our investigation of the mutagenic role of outdoor air pollution relied on an average country- and state/province-level quantification of PM_2.5_ that did not provide granularity within these geographical regions or consideration for seasonal variabilities or indoor air pollution levels. The exposure was linked to the subjects’ residence at the time of lung cancer diagnosis and may not reflect their lifetime exposure, and the assigned exposures for the patients recruited to this study may systematically differ from the average country, state, province, or city level pollution levels. As such, these associations should be interpreted with caution. Despite these limitations, our findings suggest that LCINS in individuals from areas with elevated air pollution have a higher prevalence of *TP53* mutations while exhibiting shorter telomeres and heightened mutagenesis, notably attributed to signatures SBS4 and SBS5. Consistent with our findings, a previous study comparing Chinese regions with different levels of air pollution reported an increase in *TP53* and total somatic mutations in the highly polluted region of Xuanwei, with a C>A enriched mutational pattern resembling signature SBS4^51^.

In principle, mutations linked to clock-like signature SBS5 that accumulate in LCINS with the increase of PM_2.5_ could be due to additional DNA damage that accumulates in people living in more polluted areas. In addition, considering the significant telomere shortening observed with high levels of PM_2.5_ exposure, the clock-like somatic mutations could also be a readout of a promotion mechanism where lung cells are undergoing more cell divisions in individuals residing in highly polluted areas. Consistently, a recent experimental study^48^ showed that air pollution acts as a tumor-promoting inflammatory agent, demonstrating that short-term exposure to PM_2.5_ is not strongly mutagenic in mice, although leading to an elevation of signature SBS5 and reduced telomeres (**Extended Data Fig. 11**). Future studies with long-term PM_2.5_ exposures and higher number of replicates will be required to experimentally validate the role of outdoor air pollution as a driver of both mutations and inflammation leading to tumorigenesis.

While the association between SBS4 and PM_2.5_ potentially encompasses the effects of atmospheric pollution exposure in LCINS, the small proportion of cases affected by SBS4 (6.4% of all samples) and the high number of mutations in SBS4+ cancers (21,785 on average) could indicate extreme events of outdoor, indoor, or occupational air pollution. Indeed, such events have been previously reported, for example, SBS4 was found in the lung cancers of never-smoking Chinese women that often cook with smoky coal in poorly ventilated houses^47^, and it is likely that severe levels of pollution are more common in countries with overall higher air pollution levels. Lastly, it is also possible that the self-reported smoking status was inaccurate, as previously shown in other cohorts after biochemical verification^52^, and that some SBS4 cases from smokers were misannotated as never smokers, albeit it is unlikely that such misannotation will exhibit any correlation with air pollution. Notwithstanding, the reported associations between air pollution and somatic mutations remained unaffected when all SBS4 mutations were excluded from our analysis (**Extended Data Fig. 12**).

Overall, the unprecedented size of our global cohort allowed us to refine the prevalence and intensity of the mutational signatures and driver mutations involved in LCINS, and their variability across histologies, genetic ancestries, and geographical locations. Although LCINS appear to be dominated by endogenous processes, our results indicate exposure to aristolochic acids in Taipei’s samples, a low mutagenicity of passive smoking, and a role of air pollution as both a mutagenic initiator and promoter of neoplastic expansion in LCINS.

## EXTENDED DATA FIGURE LEGENDS

**Extended Data Fig. 1. Association of mutational signature prevalence and driver mutations with geographical regions, biological sexes, and *EGFR* mutation status in LCINS adenocarcinoma cases**. **a**, DBS, ID, CN, and SV mutational signatures enrichment analysis with geographical regions. Horizontal lines marking statistically significant thresholds were included at 0.05 (dashed orange line) and 0.01 FDR value levels (dashed red line). Blue-colored signatures were enriched in North American and European patients, whereas red-colored signatures were enriched in East Asian patients. **b,** SBS, DBS, ID, CN, and SV mutational signatures enrichment analysis with biological sexes. Blue-colored signatures were enriched in males, whereas red-colored signatures were enriched in females. **c-e**, Detail of the enrichment of *EGFR* (**c**), *TP53* (**d**), and *KRAS* (**e**) driver mutations in North American and European vs. East Asian LCINS adenocarcinoma cases. **f**, Driver mutations enrichment analysis with biological sexes. Blue-colored genes were enriched in males, whereas red-colored genes were enriched in females. **h**, SBS, DBS, ID, CN, and SV mutational signatures enrichment analysis with *EGFR* mutation status. Blue-colored signatures were enriched in *EGFR* mutant tumors, whereas red-colored signatures were enriched in *EGFR* wild-type tumors.

**Extended Data Fig. 2. Tumor mutational burden differences in LCINS across histologies. a**-**b**, Quantification of tumor mutational burden according to different mutation types, including SBS, DBS, ID, number of copy number segments, and structural variants (**a**), as well as telomere length ratios between tumor and normal samples (**b**) across histologies.

**Extended Data Fig. 3. Repertoire of mutational signatures and driver mutations in LCINS carcinoids. a-b**, Mutational signature landscape for SBS (**a**) and ID (**b**) mutation types, including absolute and relative number of mutations assigned to each mutational signature, unsupervised clustering based on the signature contributions, and sample-level annotations of sex, genetic ancestry, and accuracy of signature reconstruction based on cosine similarity. **c**, Driver mutations landscape, including different types of genomic alterations, as well as sample-level annotations of sex, genetic ancestry, histology, and tumor purity.

**Extended Data Fig. 4. Association of mutational signature prevalence and driver mutations with adenocarcinoma and carcinoid histology in LCINS cases**. **a**-**b** SBS, DBS, ID, CN, and SV mutational signatures, and driver mutations (**b**) enrichment analysis. Horizontal lines marking statistically significant thresholds were included at 0.05 (dashed orange line) and 0.01 FDR value levels (dashed red line). Blue-colored signatures/genes were enriched in adenocarcinomas, whereas green-colored signatures/genes were enriched in carcinoids. **c-d**, Detail of the enrichment of *TP53* (**c**) and *ARID1A* (**d**) driver mutations in carcinoid vs. adenocarcinoma LCINS.

**Extended Data Fig. 5. Genomic landscape in LCINS squamous cell carcinomas. a-c**, Mutational signature landscape for SBS (**a**), DBS (**b**), and ID (**c**) mutation types, including absolute and relative number of mutations assigned to each mutational signature, unsupervised clustering based on the signature contributions, and sample-level annotations of sex, genetic ancestry, and accuracy of signature reconstruction based on cosine similarity. **d**, Driver mutations landscape, including different types of genomic alterations, as well as sample-level annotations of sex, genetic ancestry, histology, and tumor purity.

**Extended Data Fig. 6. Association of mutational signature prevalence and driver mutations with adenocarcinoma and squamous cell carcinoma histology in LCINS cases**. **a**-**b** SBS, DBS, ID, CN, and SV (**a**) mutational signatures, and driver mutations (**b**) enrichment analysis. Horizontal lines marking statistically significant thresholds were included at 0.05 (dashed orange line) and 0.01 FDR value levels (dashed red line). Blue-colored signatures/genes were enriched in adenocarcinomas, whereas red-colored signatures/genes were enriched in squamous cell carcinomas. **c-f**, Detail of the enrichment of *TP53* (**c**), *EGFR* (**d**), *LRP1B* (**e**), *PTEN* (**f**) and *PIK3CA* (**g**) driver mutations in squamous cell carcinoma vs. adenocarcinoma LCINS.

**Extended Data Fig. 7. Genomic landscape of 56 LCINS tumors presenting SBS4 activity. a**, Tumor mutational burden differences between SBS4 positive and negative LCINS tumors for SBS, DBS, ID, CN segments, and SV events. **b-d**, Mutational signature landscape for SBS (**b**), DBS (**c**), and ID (**d**) mutation types, including absolute and relative number of mutations assigned to each mutational signature, unsupervised clustering based on the signature contributions, and sample-level annotations of sex, genetic ancestry, passive smoking, and accuracy of signature reconstruction based on cosine similarity. **e**, Driver mutations landscape, including different types of genomic alterations, as well as sample-level annotations of sex, genetic ancestry, histology, and tumor purity.

**Extended Data Fig. 8. Topographical characteristics of 56 LCINS and 68 lung cancer from smokers presenting SBS4 activity. a-b**, Distribution of SBS4 mutations with replication timing in our cohort of never smokers (**a**) and in the smokers from the PCAWG cohort (**b**). Data are separated into deciles, with each segment harboring 10% of the observed replication time signal in the x-axis, and the normalized mutational density displayed in the y-axis. **c-d**, Association of SBS4 mutations with nucleosome occupancy in never smokers (**c**) and smokers (**d**). The solid blue line represents real somatic mutations, whereas the dashed grey line indicates the distribution of simulated mutations. Both lines show the average nucleosome signal in the y-axis, using a genomic window of 2 kilobases centered around the SBS4-associated mutations in the x-axis. **e-f**, Strand asymmetry of SBS4-associated mutations in comparison to simulations and considering lagging and leading DNA strands, transcribed and untranscribed DNA regions and genic and intergenic genomic locations in never smokers (**e**) and smokers (**f**).

**Extended Data Fig. 9. Passive smoking influence in the landscape of ID, DBS, CN, and SV in LCINS. a-e**, Differences in DBS, ID, CN, and SV burden using univariate comparisons (**a**) as well as multivariable linear regressions considering clinical and epidemiological covariates (**b-e**), including age, sex, genetic ancestry, and tumor purity. **f**, Volcano plots indicating enrichment of mutational signatures derived from DBS, ID, CN, and SV alterations. Horizontal lines marking statistically significant thresholds were included at 0.05 (dashed orange line) and 0.01 FDR value levels (dashed red line).

**Extended Data Fig. 10. Effects of PM_2.5_ exposure in large genomic alterations in LCINS. a-b**, Differences in the number of CN segments and SV events using univariate comparisons (**a**) as well as multivariable linear regressions, considering clinical and epidemiological covariates (**b**), including age, sex, genetic ancestry, histology, and tumor purity, for patients diagnosed in geographical regions with high and low PM_2.5_ exposure levels (threshold defined at 20 μg/m^3^; only samples for which the country of origin was known, *n*=853, were included). **c**, Volcano plots indicating enrichment of mutational signatures derived from CN and SV alterations. Horizontal lines marking statistically significant thresholds were included at 0.05 (dashed orange line) and 0.01 FDR value levels (dashed red line).

**Extended Data Fig. 11. Assignment of mutational signatures and estimation of telomere length using data from control (*n*=5) and PM_2.5_-exposed mice (*n*=5) from Ref.^48^. a-b**, Boxplots comparing the mutations assigned to SBS5 (**a**) and the estimations for the telomere length ratio between the tumor and normal samples (**b**). Student’s t-tests were used to calculate statistical significance.

**Extended Data Fig. 12. Mutagenic effects of PM_2.5_ exposure in LCINS cases excluding SBS4 contributions. a**, Quantification of SBS burden excluding SBS4 mutations for patients living in geographical regions with high and low PM_2.5_ exposure levels (threshold defined at 20 μg/m^3^; only samples for which the country of origin was known, *n*=853, were included). **b**, Forest plot corresponding to a multivariable linear regression considering high/low PM_2.5_ exposure group, age, sex, genetic ancestry, histology, and tumor sample purity as covariates and SBS burden as independent variable. **c**, Scatter plot showing a significant correlation between individual sample estimates of PM_2.5_ exposure and SBS burden.

**SUPPLEMENTARY MATERIAL LEGENDS**

**Supplementary Table 1. Clinical characteristics of the Sherlock-*Lung* never-smoking lung cancer cohort by histology.**

**Supplementary Table 2. Mutational profiles for SBS *de novo* extracted mutational signatures using the SBS-288 mutational context.**

**Supplementary Table 3. Mutational profiles for ID *de novo* extracted mutational signatures using the ID-83 mutational context.**

**Supplementary Table 4. Mutational profiles for DBS *de novo* extracted mutational signatures using the DBS-78 mutational context.**

**Supplementary Table 5. Mutational profiles for CN *de novo* extracted mutational signatures using the CN-68 mutational context.**

**Supplementary Table 6. Mutational profiles for SV *de novo* extracted mutational signatures using the SV-38 mutational context.**

**Supplementary Table 7. Decomposition of SBS *de novo* mutational signatures into COSMICv3.4 reference signatures.**

**Supplementary Table 8. Decomposition of ID *de novo* mutational signatures into COSMICv3.4 reference signatures.**

**Supplementary Table 9. Decomposition of DBS *de novo* mutational signatures into COSMICv3.4 reference signatures.**

**Supplementary Table 10. Decomposition of CN *de novo* mutational signatures into COSMICv3.4 reference signatures.**

**Supplementary Table 11. Decomposition of SV *de novo* mutational signatures into COSMICv3.4 reference signatures.**

**Supplementary Table 12. Associations of passive smoking and pollution with genomic features adjusted by genetic ancestry principal components.**

**Supplementary Table 13. dbGaP and EGA unique identifiers for the publicly available datasets included as part of the analyzed LCINS cohort.**

**Supplementary Fig. 1. *De novo* mutational signatures extracted using the SBS-288 mutational context**. **a**, Mutational profiles of the *de novo* extracted signatures, with indication of the cosine similarity of the decomposition into COSMICv3.4 reference signatures. **b**, Contribution of the different COSMICv3.4 reference mutational signatures after decomposition of the *de novo* extracted signatures. **c**, Activity of *de novo* mutational signatures across samples, representing the total number of substitutions attributed to each signature in a given sample. Dots represent individual samples, colors different histology types, and purple horizontal bars median values across all histologies. The numbers on top indicate the total number of samples where a particular signature was found active (blue) and the total number of samples of the assessed cohort (green).

**Supplementary Fig. 2. *De novo* mutational signatures extracted using the ID-83 mutational context**. **a**, Mutational profiles of the *de novo* extracted signatures. **b**, Contribution of the different COSMICv3.4 reference mutational signatures after decomposition of the *de novo* extracted signatures. **c**, Activity of *de novo* mutational signatures across samples, representing the total number of indels attributed to each signature in a given sample. Dots represent individual samples, colors represent different histology types, and purple horizontal bars represent median values across all histologies. The numbers on top indicate the total number of samples where a particular signature was found active (blue) and the total number of samples of the assessed cohort (green).

**Supplementary Fig. 3. *De novo* mutational signatures extracted using the DBS-78 mutational context**. **a**, Mutational profiles of the *de novo* extracted signatures. **b**, Contribution of the different COSMIC v3.4 reference mutational signatures after decomposition of the *de novo* extracted signatures. **c**, Activity of *de novo* mutational signatures across samples, representing the total number of doublets attributed to each signature in a given sample. Dots represent individual samples, colors represent different histology types, and purple horizontal bars represent median values across all histologies. The numbers on top indicate the total number of samples where a particular signature was found active (blue) and the total number of samples of the assessed cohort (green).

**Supplementary Fig. 4. *De novo* mutational signatures extracted using the CN-68 mutational context**. **a**, Mutational profiles of the *de novo* extracted signatures. **b**, Contribution of the different COSMICv3.4 reference mutational signatures after decomposition of the *de novo* extracted signatures. **c**, Activity of *de novo* mutational signatures across samples, representing the total number of copy number segments attributed to each signature in a given sample. Dots represent individual samples, colors represent different histology types, and purple horizontal bars represent median values across all histologies. The numbers on top indicate the total number of samples where a particular signature was found active (blue) and the total number of samples of the assessed cohort (green).

**Supplementary Fig. 5. *De novo* mutational signatures extracted using the SV-38 mutational context**. **a**, Mutational profiles of the *de novo* extracted signatures. **b**, Contribution of the different COSMIC v3.4 reference mutational signatures after decomposition of the *de novo* extracted signatures. **c**, Activity of *de novo* mutational signatures across samples, representing the total number of structural variants attributed to each signature in a given sample. Dots represent individual samples, colors represent different histology types, and purple horizontal bars represent median values across all histologies. The numbers on top indicate the total number of samples where a particular signature was found active (blue) and the total number of samples of the assessed cohort (green).

**Supplementary Fig. 6. Validation of homologous recombination deficient cases in LCINS using computational predictors**. The x-axis shows the probability prediction scores for all the LCINS cases in the cohort (categorized by histology and presence of signature SBS3) using HRDetect^31^, whereas the y-axis shows the probability prediction scores for CHORD^30^. Dashed lines showed the thresholds proposed by both computational tools for considering a sample as homologous recombination deficient or proficient.

**Supplementary Fig. 7. Sensitivity analysis of the associations of mutational signatures with geographical regions excluding Canadian patients with EAS genetic ancestry.** Volcano plot indicating enrichment of SBS signatures in patients from East Asian (AS) and North American/European regions (NA/EU) in LCINS adenocarcinomas (top panel) and bar plot indicating prevalence by geographical region (bottom panel). Horizontal lines marking statistically significant thresholds were included at 0.05 (dashed orange line) and 0.01 FDR levels (dashed red line).

**Supplementary Fig. 8. Passive smoking influence in the genomic landscape of lung adenocarcinomas from never smokers. a-d**, Differences in base substitution burden and telomere length ratio between tumor and normal samples using univariate comparisons (**a** and **b**) as well as multivariable linear regressions considering clinical and epidemiological covariates (**c** and **d**), including age, sex, genetic ancestry, and tumor purity.

**Supplementary Fig. 9. Assessment of the resolution to assign signature SBS4 in cases exposed to secondhand smoke.** COSMICv3.4 mutational signatures obtained for the whole cohort of LCINS patients were assigned to 247 tumors from SBS4-negative patients exposed to secondhand smoke after the synthetic injection of SBS4 at different levels (1%, 2%, 5%, 10%, 15%, and 20%; 100 simulations per injection level). **a**, Boxplots showing the distribution of the proportion of samples where SBS4 was detected for different simulations within a synthetic injection level. **b**, Detail of the SBS4 contributions observed in the synthetic samples, where each row corresponds to a sample, each column to a simulation corresponding to a specific injection level, and the color represents the contribution level of SBS4 to the overall mutational profile.

**Supplementary Fig. 10. Split of samples in pollution exposure groups according to their levels of estimated population weighted PM_2.5_.** Limit used for the two-group classification (high-low; 20 μg/m^3^; 440 high exposed samples vs. 413 low exposed samples).

## ONLINE METHODS

### Ethics declarations

Since the National Cancer Institute only received de-identified samples and data from collaborating centers, had no direct contact or interaction with the study participants, and did not use or generate identifiable private information, Sherlock-*Lung* has been determined to constitute “Not Human Subject Research (NHSR)” based on the federal Common Rule (45 CFR 46; https://www.ecfr.gov/cgi-bin/ECFR?page=browse).

### Collection of lung cancer samples

Fresh-frozen tumor tissue and matched germline DNA from whole-blood samples or fresh-frozen normal lung tissue sampled approximately 3 cm from the tumor were obtained from 871 treatment-naïve lung cancer patients from 14 institutions/centers across the world after sample and sequencing quality control. Among these patients, 114 were from the Institut universitaire de cardiologie et de pneumologie de Québec – Université Laval (IUCPQ-UL), Quebec, Canada; 54 from Université Côte d’Azur, Nice, France; 25 from the EAGLE study from Italy^53^; 22 from Yale University, New Haven, Connecticut, USA; 11 from H. Lee Moffitt Cancer Center & Research Institute, Tampa, Florida, USA; 27 from Harvard University, Cambridge, Massachusetts, USA; 113 from Hong-Kong; 192 from the International Agency for Research on Cancer, Lyon, France; 13 from Mayo Clinic, Rochester, Minnesota, USA; 13 from Roswell Park Comprehensive Cancer Center, Buffalo, New York, USA; 185 from Taipei; 68 from Toronto, Canada; 5 from Valencia, Spain; and 29 from previously published and publicly available whole-genome sequenced (WGS) lung cancers from never smokers (**Supplementary Table 13**). For these 871 individuals, the mean age at lung cancer diagnosis was 64.1 years (range: 21–92) and 79.0% of patients were female.

Similar to our prior publication^19^, we utilized seven rigorous criteria for sample inclusion: *(i) Sequencing coverage.* We maintained a minimum average sequencing coverage of >40x for tumor samples and >25x for normal samples; *(ii) Contamination and Relatedness.* Cross-sample contamination was limited to <1% by Conpair^54^, and detected relatedness was maintained <0.2 by Somalier^55^; *(iii) Copy number analysis.* Subjects with abnormal copy number profiles in normal samples were excluded, as determined by Battenberg^56^; *(iv) Mutational signatures.* Tumor samples exhibiting mutational signatures SBS7 (associated with ultraviolet light exposure^41^) and SBS31 (associated with platinum chemotherapy^57^) were removed from the analysis. *(v) Tumor type validation.* Tumor samples reported as non-lung cancer or not originating from primary lung cancer were excluded. *(vi) WGS quality control.* Tumor samples with a total genomic alteration count of <100 or <1000 along with NRPCC (the number of reads per clonal copy)^58^ <10 were excluded as low-quality samples. *(vii) Multiple-region sequencing.* In rare cases where multiple regions of a tumor were sequenced, only one high-purity tumor sample was included to avoid redundancy. These stringent criteria were applied consistently to ensure the robustness and reliability of the data collected for the Sherlock-*Lung* study.

All 871 matched tumor and germline samples underwent DNA WGS. Except for the 29 previously generated WGS cancers^18,59-62^, sequencing was performed the same for all other samples. Specifically, frozen tumor tissue with matched blood or normal tissue samples were immediately put into 1ml of 0.2 mg/ml Proteinase K (Qiagen) in DNA lysis buffer (10 mMTris-Cl (pH 8.0), 0.1 M EDTA (pH 8.0), and 0.5% (w/v) SDS) for 24 hours at 56°C with shaking at 850 rpm in Thermomixer R (Eppendorf) until the tissue was completely lysed. Genomic DNA was extracted from fresh frozen tissue using the QIAmp DNA Mini Kit (Qiagen) according to the manufacturer’s instructions. Each sample was eluted in 200 μl AE buffer and DNA concentration was determined by Nanodrop spectrophotometer. All DNA samples were aliquoted and stored at −80°C until use.

DNA was quantified using the QuantiFluor® dsDNA System (Promega Corporation, USA). DNA was normalized to 25ng/ul and underwent fragment analysis via AmpFLSTR™ Identifiler™ PCR Amplification Kit (ThermoFisher Scientific, USA). DNA samples are required to meet minimum mass and concentration thresholds for each assay, as well as show no evidence of contamination or profile discordance in the Identifiler assay. Samples meeting these requirements were aliquoted at the appropriate mass needed for downstream assay processing.

### Whole-genome sequencing (WGS)

The resulting post-capture enriched multiplexed sequencing libraries were used in cluster formation on an Illumina cBOT (Illumina, San Diego, CA, USA). Paired-end sequencing was performed by the Broad Institute (https://www.broadinstitute.org) using the Illumina HiSeq X system following Illumina-provided protocols for 2x151bp paired-end sequencing. FASTQ files were generated after Illumina base-calling. Next, paired FASTQ were converted to unmapped BAM using the GATK pipeline (https://github.com/gatk-workflows/seq-format-conversion). The unmapped BAM files were then processed using GATK on the cloud-based platform TERRA workspaces (https://app.terra.bio). The sequence data were aligned to the human reference genome GRCh38, and the aligned BAM files were transferred to the NIH HPC system (https://hpc.nih.gov) for downstream analyses.

For the public WGS data, the preprocessed aligned BAM/CRAM files (including unmapped reads) were first converted back to FASTQ files using Bazam (v.1.0.1)^63^ to retain the sequencing lane and read group information and then processed using the same pipeline as for the Sherlock-*Lung* WGS dataset.

### Somatic variant calling

The somatic variant calling was performed using our established bioinformatics pipeline as previously described^19^. The analysis-ready BAM files were processed using four different mutation calling algorithms for tumor-normal paired analysis, including MuTect^64^, MuTect2, Strelka v.2.9.10^65^ and TNscope^66^, implemented in the Sentieon’s genomics software (v202010.01). We employed an ensemble method to merge the results from these different callers followed by additional filtering to reduce false positive calling. The final mutation calls for both single base substitutions (SBSs) and small insertions and deletions (indels) were required to meet the following criteria: *(i)* read depth >12 in tumor samples and >6 in normal samples; *(ii)* variant allele frequency <0.02 in the matched-normal sample; and *(iii)* overall allele frequency (AF) <0.001 in multiple genetics databases including 1000 Genomes (phase 3 v5), gnomAD exomes (v2.1.1), and gnomAD genomes (v3.0)^67^. The filtered variants were annotated with Oncotator v.1.9.1.0^68^ and ANNOVAR v.2019-10-2495. For the indel calling, only variants called by at least three algorithms were kept (MuTect2, Strelka, and TNscope). The UPS-indel^69^ algorithm was used to compare and combine different indel call sets. Similar filtering steps as those used for SNV calling were also applied to indel calling. The final set of indels were left-normalized (left-aligned and trimmed) for the downstream analysis.

### Ancestry estimation

To confirm the genetic ancestry of the patients, we calculated the principal component (PC) coordinates based on WGS data. This analysis was performed using the VerifyBamID (v.2.0.1) algorithm^70^ in conjunction with samples from the 1000 Genomes Project^21^.

### Driver gene discovery

The IntOGen pipeline v2020.02.0123^71^, which combines seven state-of-the-art computational methods, was employed to detect signals of positive selection in the mutational pattern of driver genes across the cohort. The 65 genes identified as drivers with combination q-value<0.1 in the cohort were classified according to their mode of action in tumorigenesis (*i.e.*, tumor suppressor genes or oncogenes) based on the relationship between the excess of observed nonsynonymous and truncating mutations computed by dNdScv^72^ and their annotations in the Cancer Gene Census^73^. Genes with conflicting computed and annotated modes of action were labeled ambiguous. To identify potential driver mutations across the 65 cancer driver genes annotated in the Cancer Gene Census, we selected mutations that fulfilled any of the following criteria: *(i)* truncating mutations in genes annotated as tumor suppressors; *(ii)* recurrent missense mutations (seen in at least three independent tumors); *(iii)* mutations classified as “Likely Drivers” by boostDM (score >0.5)^74^; *(iv)* mutations classified as “Oncogenic” or “Likely Oncogenic” by OncoKB^75^; *(v)* mutations classified as drivers in the TCGA MC3 drivers study^76^; *(vi)* missense mutations classified as “Likely Pathogenic” by AlphaMissense^77^ in genes annotated as tumor suppressors.

### Estimation of tumor purity, ploidy, and allele-specific copy numbers

We used the Battenberg algorithm (v2.2.9)^56^ to conduct analyses of somatic copy number alterations (SCNA). Initial SCNA profiles were generated, followed by an assessment of the clonality of each segment, purity, and ploidy. Any SCNA profile determined to have low-quality after manual inspection underwent a refitting process using the Battenberg algorithm. This process required new tumor purity and ploidy inputs, either estimated by ccube (v1.0)^78^ or recalculated from local copy number status. The Battenberg refitting procedures were iteratively executed until the final SCNA profile was established and met the criteria of manual validation check. GISTIC (v2.0)^79^ was used to identify the recurrent copy number alterations at the gene level based on the major clonal copy number for each segmentation.

### Structural variants calling

Meerkat (v.0.189)^80^ and Manta (v.1.6.0)^81^ were applied with recommended filtering for identifying structural variants (SVs), and the union set of these two callers was merged as the final SV dataset.

### Telomere length

We estimated telomere length in kb using TelSeq (v.0.0.2)^82^ for all 871 LCINS samples as well as mouse data from a recent experimental study, including control and PM_2.5_-exposed mice^48^. We used seven as the threshold for the number of TTAGGG/CCCTAA repeats in a read for the read to be considered telomeric. The TelSeq calculation was done individually for each read group within a sample, and the total number of reads in each read group was used as weight to calculate the average TL for each sample.

### Mutational signature analysis

#### Extraction of de novo mutational signatures

*De novo* mutational signatures for single base substitutions (SBS), doublet base substitutions (DBS), and indels (ID) were extracted using SigProfilerExtractor^22^ v1.1.21 with default parameters and normalization set to 10,000 mutations, in order to limit the effect of hypermutators in the signature extraction process. For SBSs, *de novo* signatures were extracted using the SBS-288 and SBS-1536 high-definition mutational contexts, which, beyond the common SBS-96 trinucleotide context using the mutated base and the 5’ and 3’ adjacent nucleotides^83,84^, also consider the transcriptional strand bias and the pentanucleotide context (two 5’ and 3’ adjacent nucleotides), respectively^84^. Given the high similarity obtained for both mutational contexts as well as the additional separation of mutational processes obtained by the SBS-288 mutational context (11 *de novo* signatures using SBS-288 vs. 10 *de novo* signatures using SBS-1536; average cosine similarity 0.97), the results using the SBS-288 context were used for further analysis (**Supplementary Table 2**). Previously established mutational contexts DBS-78 and ID-83^15,84^ were used for the extraction of DBS and ID signatures (**Supplementary Tables 3-4**). Copy number signatures were extracted *de novo* following an updated context definition benefitting from deep WGS data (CN-68) (**Supplementary Table 5**), which allowed to further characterize CN segments below 100kbp in length (in contrast to current COSMICv3.4 reference signatures using the CN-48 context, which are based on SNP6 microarray data and therefore without the resolution to characterize short CN segments)^26^. SV signatures were extracted using a similarly refined context, with an in-depth characterization of short SV alterations below 1kbp (SV-38 context; **Supplementary Table 6**).

#### Decomposition and assignment of mutational signatures to individual tumors

After *de novo* extraction was completed, SigProfilerAssignment^85^ v0.1.1 was used to decompose the *de novo* extracted SBS, ID, DBS, CN, and SV mutational signatures into COSMICv3.4^23^ reference signatures based on the GRCh38 reference genome (**Supplementary Tables 7-11**) as well as to assign signatures to individual samples obtaining signature activities, based on the forward stagewise algorithm for sparse regression and nonnegative least squares for numerical optimization. Hierarchical clustering of the activities of mutational signatures was performed using Euclidean distance and Ward’s minimum-variance clustering. For the SBS signatures, the 11 *de novo* extracted signatures were originally decomposed into 15 COSMICv3.4 reference signatures. However, two of the COSMICv3.4 signatures, enriched in C>T substitutions, were removed from the decomposition to avoid misassignment, namely SBS23 and SBS32, as the patterns of strong transcriptional strand bias generated by these two signatures^15^ were not observed in our LCINS dataset, and their individual contribution to the decomposition of *de novo* signatures was minimal. On the other hand, five additional COSMICv3.4 SBS signatures were included for the assignment to individual samples, including SBS3, SBS21, SBS33, SBS44, and SBS92. SBS3 and SBS92 were included considering their previously reported strong associations with indel signatures ID6 and ID3, both of which were observed in our indel signature analysis after decomposition to COSMICv3.4 ID signatures^15,22,43^. Thus, for those tumors harboring ID6 (*n*=41), we allowed assigning SBS3 (assigned only to 14 of the cases), whereas for those tumors with presence of ID3 (*n*=300), SBS92 was allowed in the assignment (assigned to 6 tumors). Suspected homologous recombination deficient (HRD) tumors exhibiting signatures SBS3 and ID6 were tested using two independent computational algorithms, CHORD^30^ and HRDetect^31^. Lastly, two samples showing a low cosine similarity for the mutational profile reconstruction (NSLC-0477 and NSLC-0637) showed high similarities in their mutational profiles with signatures linked to microsatellite instability (MSI) and COSMIC SBS33, respectively. Considering this, we allowed assignment of SBS33 and all MSI-associated signatures (SBS6, SBS14, SBS15, SBS20, SBS21, SBS26, and SBS44), respectively, in each of this specific samples, with SBS21 and SBS44 being finally assigned to sample NSLC-0477. The MSI phenotype of this sample was independently confirmed by MMRDetect^25^, and further evidence was collected by exploring the indel landscape, with 10,474 alterations, 29-fold more than the average number of indels in lung adenocarcinomas, and a strong enrichment of signature ID2, consistent with previously reported MSI cases^15^. The decomposed COSMICv3.4 SBS signatures obtained in the whole LCINS cohort were used for the assignment of signatures to the mouse data obtained from a recent publication^48^, after their renormalization to the mouse genome (mm10 build).

#### Assignment of mutational signatures to individual somatic mutations

Signatures were probabilistically assigned to individual somatic mutations using SigProfilerAssignment based on Bayes’ rule and the specific mutational context for the mutation, as previously described^85^. Briefly, to calculate the probability of a specific mutational signature being responsible for a mutation in a given mutational context and in a particular sample, we multiplied the general probability of the signature causing mutations in a specific mutational context (obtained from the mutational signature profile) by the activity of the signature in the sample (obtained from the signature activities), and then normalized this value dividing by the total number of mutations corresponding to the specific mutational context (obtained from the reconstructed mutational profile of the sample).

### Assessment of statistical power to assign mutational signatures in passive smokers

To assess the statistical power to assign SBS4 in passive smokers, we performed simulations where SBS4 was injected at different average levels (1%, 2%, 5%, 10%, 15%, and 20% of total mutations in each sample; 100 simulations for each injection level) in all 247 tumors from passive smoker patients lacking SBS4. For each sample, prior to the injection of SBS4, the number of mutations to be injected into the sample was randomly subtracted from the sample while ensuring all mutation counts were still non-negative. Next, SBS4 mutations were injected at the current level being tested. After, 10% Gaussian noise was added to the resulting mutational profile. Subsequently, mutational signatures were re-assigned in each sample using SigProfilerAssignment^85^ as well as the 18 SBS COSMICv3.4 reference signatures considered for the original data using default parameters except for a relaxed addition penalty (nnls_add_penalty=0.01) in order to increase the sensitivity of the signature assignment analysis.

### Evaluating the topography of mutational signatures

Topography analyses specific for the 56 SBS4 positive samples as well as 68 lung cancer samples from smoker patients from the Pan-Cancer Analysis of Whole Genomes (PCAWG) cohort^45^ were carried out with SigProfilerTopography^86^, which evaluates the effect of DNA replication, DNA transcription, chromatin organization, histone modifications, and transcription factor binding on the activities of different mutational processes. SigProfilerTopography examines the distribution of topographical features and narrows down the analyses by calculating the average signal of each feature in the close vicinity of the somatic mutations. Next, all the results of somatic mutations are statistically compared with those from simulated mutations that account for the patterns of all operative mutational signatures within an examined sample to elucidate statistically significant differences.

### Estimating air pollution

Annual country-level population-weighted mean concentration estimates of the environmental particulate matter measuring ≤2.5 μm (PM_2.5_) (μg/m^3^) from 1998–2021 were obtained from a hybrid model combining measurements of aerosol optical depth from different satellites (MODIS, VIIRS, MISR, and SeaWiFS) with chemical transport modeling (GEOS-Chem) and ground-based photometer (AERONET) observations^49^, and downloaded from https://sites.wustl.edu/acag/datasets/surface-pm2-5/. Additional state-level and provincial-level yearly mean estimates were also used for the patients from the United States and Canada, respectively. An individual estimate of the PM_2.5_ exposure for each patient was calculated by averaging the annual mean concentration values from the year of lung cancer diagnosis (calculated by adding the age of diagnosis to the year of birth) until the earliest year with data available, i.e., 1998. For two patients diagnosed before 1998, as well as for 28 patients whose age of diagnosis or year of birth was unknown, the PM_2.5_ estimates for 1998 were considered. PM_2.5_ estimates were not available for 18 cases for which the country of residence was unknown. For the dichotomized analysis between high and low polluted regions according to the levels of individual estimates of the PM_2.5_ exposure, a threshold of 20 μg/m^3^ was used, considering the distribution of PM_2.5_ estimates across the whole cohort (**Supplementary Fig. 10**).

### Association of geographical regions, histologies, *EGFR* driver mutation status, air pollution, and passive smoking with genomic features

In order to quantify the influence of passive smoking and air pollution on the number of total somatic mutations and the ratio of telomere length between tumor and normal samples, we fitted a series of multivariable linear models, including adjustments for several confounding covariates, namely age, sex, genetic ancestry, histology, and tumor purity. The total number of mutations (log_10_ scale) or telomere length ratio (log_2_ scale) served as dependent variables for the linear regressions, respectively, whereas passive smoking/air pollution groups and covariates were used as independent variables. In addition, for assessing the dose-response effect of air pollution, we fitted multivariable linear regressions using the individual PM_2.5_ estimates per sample, considering similar adjustments as in the analysis by pollution groups. Genetic ancestry was primarily considered for the adjustments based on the super-samples from the 1,000 Genomes Project^21^, classifying the samples into EUR, EAS, or Other genetic ancestry (**Fig. 1*b***). Additionally, we also considered the genetic ancestry principal components derived from the WGS data for the adjustments, with no differences in the associations compared to the ancestry label-based adjustments (**Supplementary Table 12**)

Similarly, the presence of specific mutational signatures or mutations in specific driver genes was modeled based on the influence of geographical region/histology/*EGFR* mutation status/passive smoking/air pollution and the additional covariates previously considered, including age, sex, genetic ancestry, tumor purity, and histology (if applicable), by using multivariable logistic regressions. For this purpose, signature activities were dichotomized into the presence or absence of a particular signature, and these binary activities served as dependent variables for the logistic regressions, with geographical regions, *EGFR* mutation status, histology subtypes, passive smoking groups, or individual PM_2.5_ estimates, respectively, along with covariates, being used as independent variables. For signatures present in more than 50% of the cases, dichotomization was done above and below the median of assigned mutations to the overall cohort of LCINS patients (*n*=871). Signatures SBS21, SBS33, and SBS44 were excluded from the analysis as they were only found in individual tumors. For the enrichment analysis of mutations in driver genes, the presence or absence of mutations in specific genes was used as the dependent variable for the logistic regressions. Only driver genes having driver mutations in more than 2% of cases of the assessed cohort were considered.

In addition, to assess the dose-response effect of air pollution, we fitted multivariable linear regressions using the individual PM_2.5_ estimates per sample, considering similar corrections for covariates and the tumor mutational burden (log_10_ scale), the telomere length ratio (log_2_ scale), or the number of mutations contributed by a given signature (log_10_ scale) as the dependent variable. Only samples where a particular mutational signature was present were considered for the multivariable linear regressions. Univariate linear regressions for the average tumor mutational burden (log_10_ scale), the telomere length ratio (log_2_ scale), or the number of mutations contributed by a given signature (log_10_ scale) vs. the average PM_2.5_ per geographical region were used for visualization.

In all cases, p-values were corrected according to the different signatures from the same variant type or driver genes considered by using a false-discovery rate correction based on the Benjamini-Hochberg method^87^ and reported as FDR. FDR<0.05 were considered statistically significant.

### Statistical analysis

All statistical analyses and graphic displays were performed using the R software v4.2.3 (https://www.r-project.org/). Two-sided Fisher’s exact tests were used for the enrichment analyses of categorical variables. For the comparison of numerical variables across groups, we used non-parametric Mann-Whitney (Wilcoxon rank sum) tests. P-values<0.05 were considered statistically significant. If multiple hypothesis testing was required, we used a false-discovery rate correction based on the Benjamini-Hochberg method^87^ and reported FDR. FDR<0.05 were considered statistically significant.

## DATA AVAILABILITY

Normal and tumor-paired CRAM files for the 871 WGS subjects of the Sherlock-*Lung* study have been deposited in dbGaP under the accession numbers phs001697.v1.p1. Detailed access information for the publicly available datasets can be found in **Supplementary Table 13**.

## CODE AVAILABILITY

The WGS bioinformatics pipelines can be accessed at https://github.com/xtmgah/Sherlock-Lung.

Battenberg SCNA calling algorithm can be found at https://github.com/Wedge-lab/battenberg.

## Data Availability

Normal and tumor-paired CRAM files for the 871 WGS subjects of the Sherlock-Lung study have been deposited in dbGaP under the accession numbers phs001697.v1.p1.

## ACKNOWLEDGEMENTS

This work was supported by the Intramural Research Program of the National Cancer Institute, US National Institute of Health (NIH) (project ZIACP101231 to MTL); by the NIH grants R01ES032547-01, R01CA269919-01, and 1U01CA290479-01 to LBA as well as by LBA’s Packard Fellowship for Science and Engineering. The research performed in LBA’s lab was also supported by UC San Diego Sanford Stem Cell Institute. The funders had no roles in study design, data collection and analysis, decision to publish, or preparation of the manuscript. The computational analyses reported in this manuscript have utilized the Triton Shared Computing Cluster at the San Diego Supercomputer Center of UC San Diego. We thank the study participants, Dr. Peter Kraft for his reviewing of the manuscript and insightful comments, and the staff at Westat Inc. for their valuable assistance in collecting samples and corresponding clinical data. This work utilized the computational resources of the NIH high-performance computational capabilities Biowulf cluster (http://hpc.nih.gov).

