## Extended Data Figures for "The mutagenic forces shaping the genomic landscape of lung cancer in never smokers"

### Extended Data Fig. 1

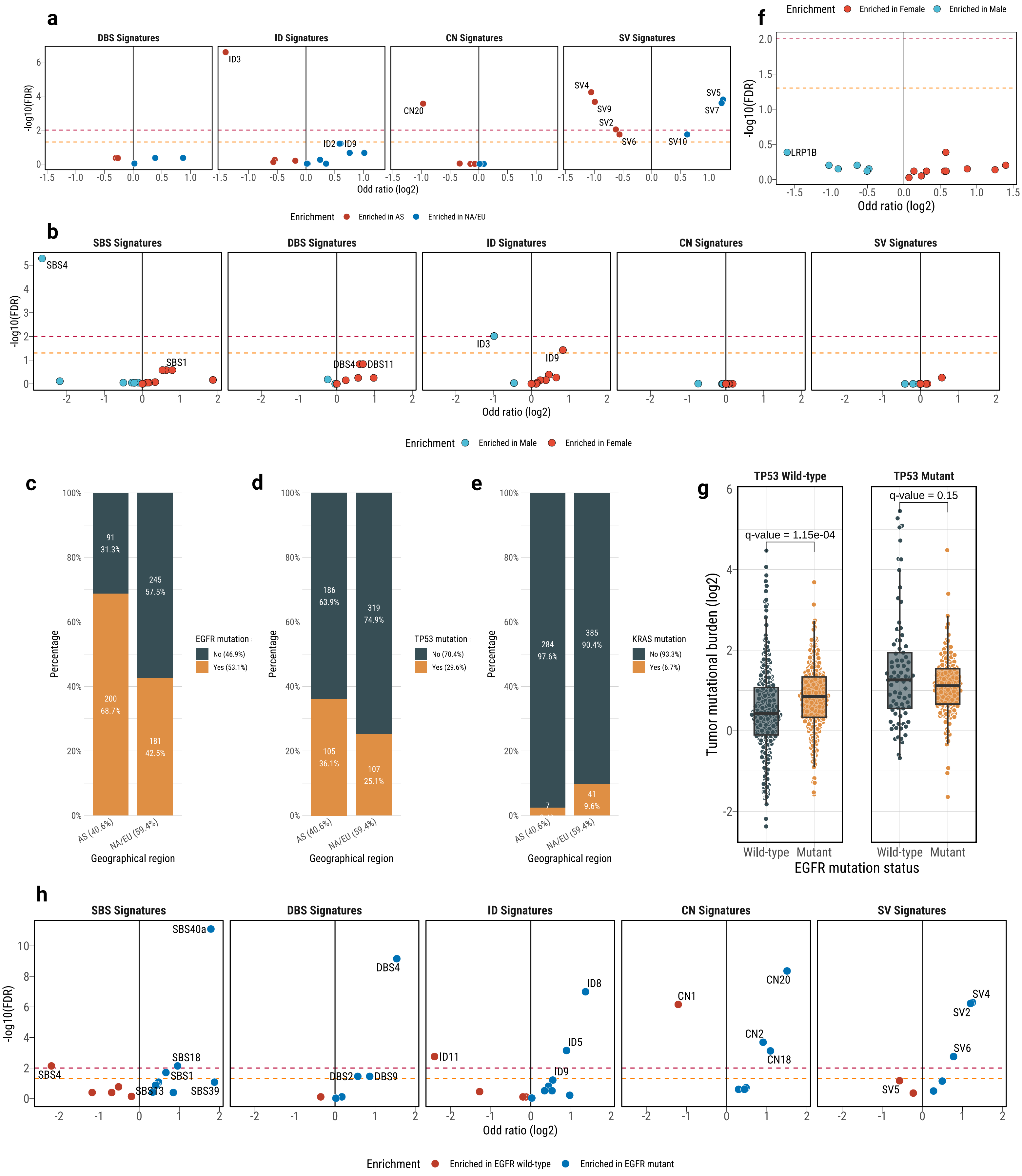

#### Extended Data Fig. 2

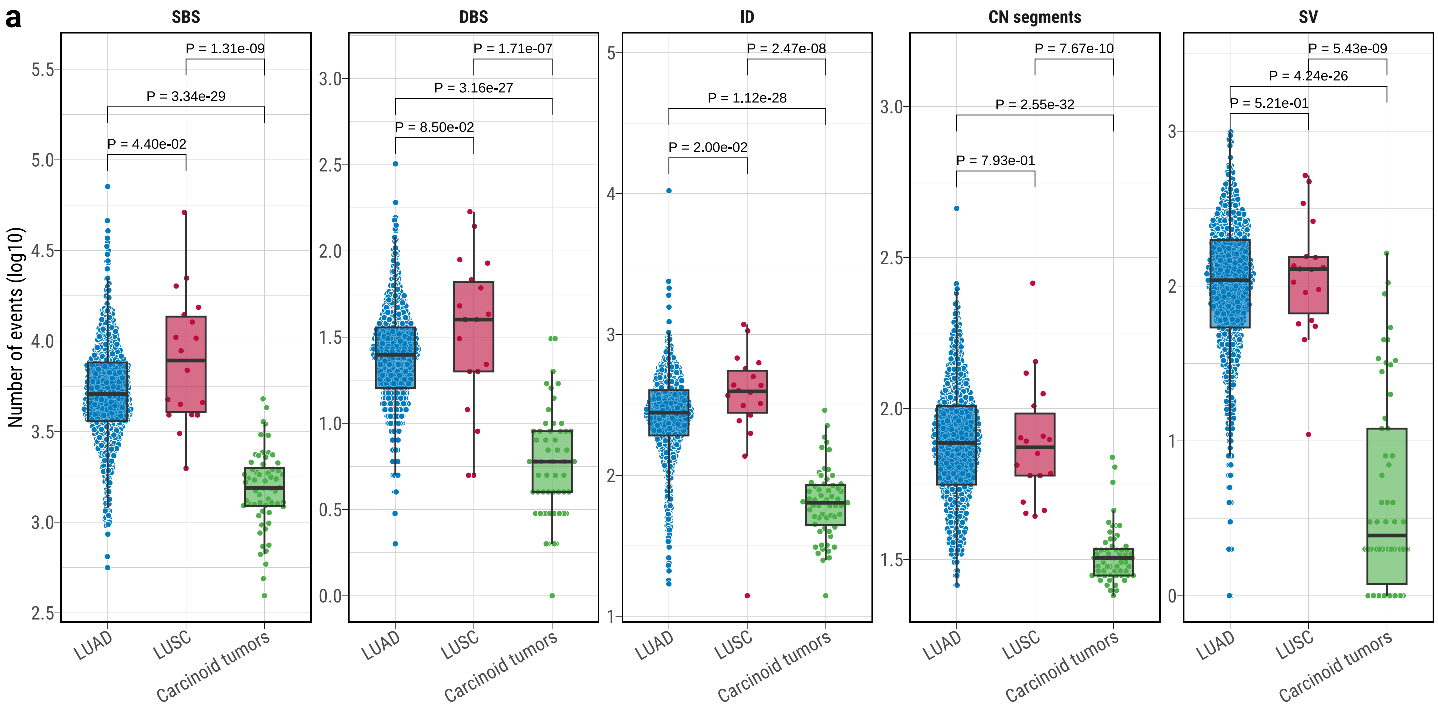


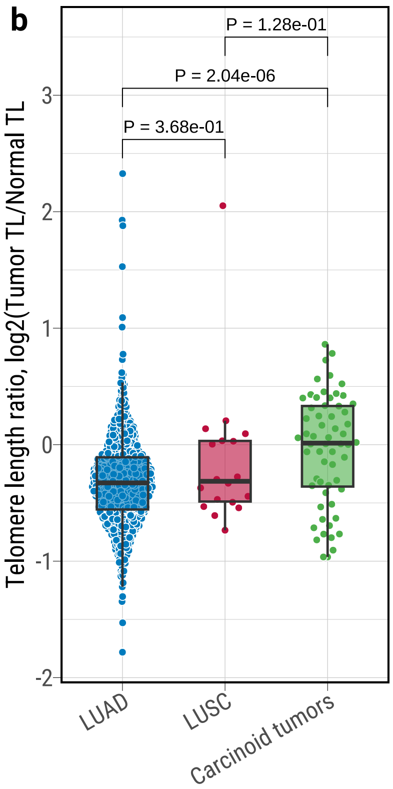

#### Extended Data Fig. 3

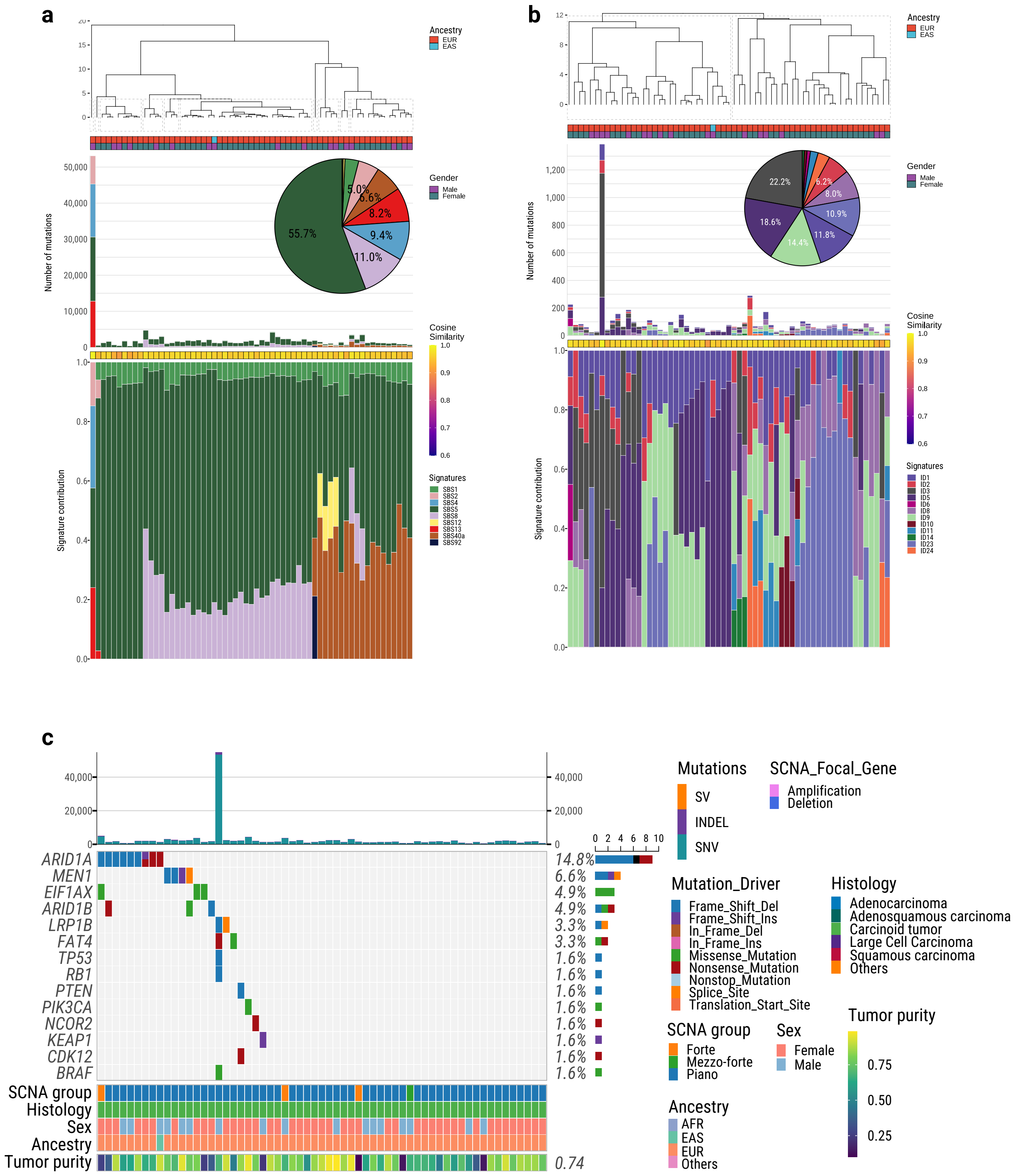

### Extended Data Fig. 4

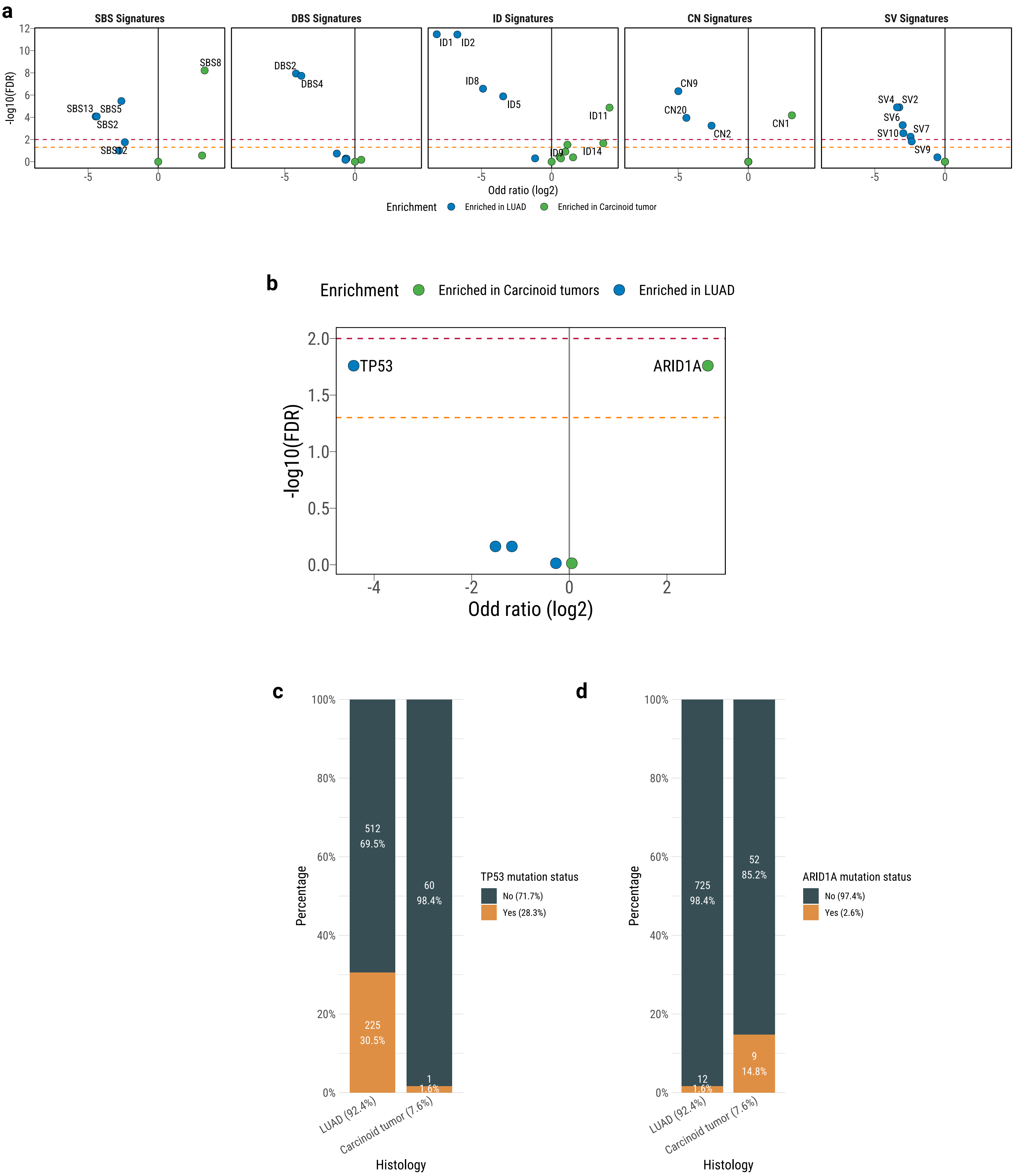

### Extended Data Fig. 5

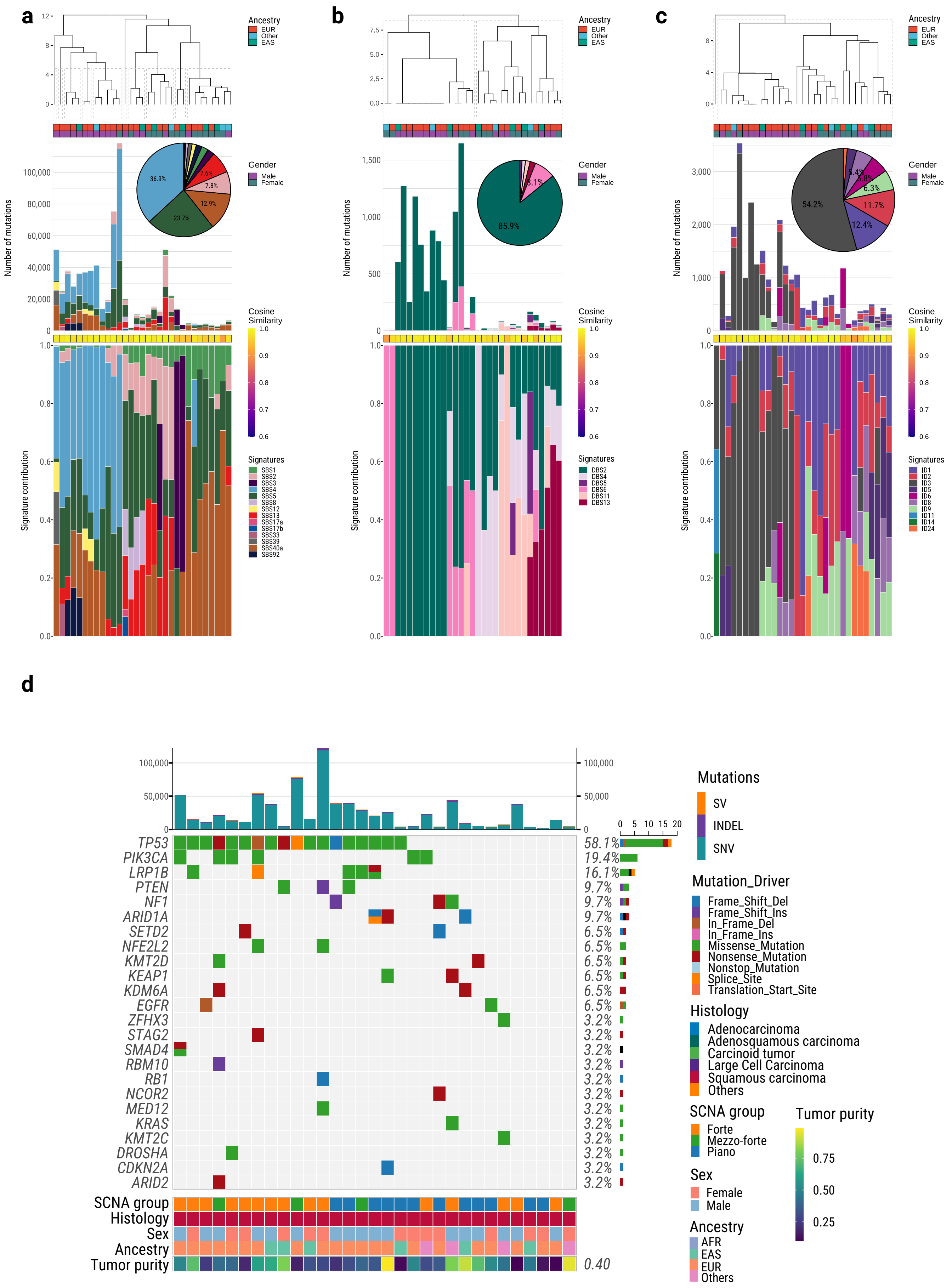

### Extended Data Fig. 6

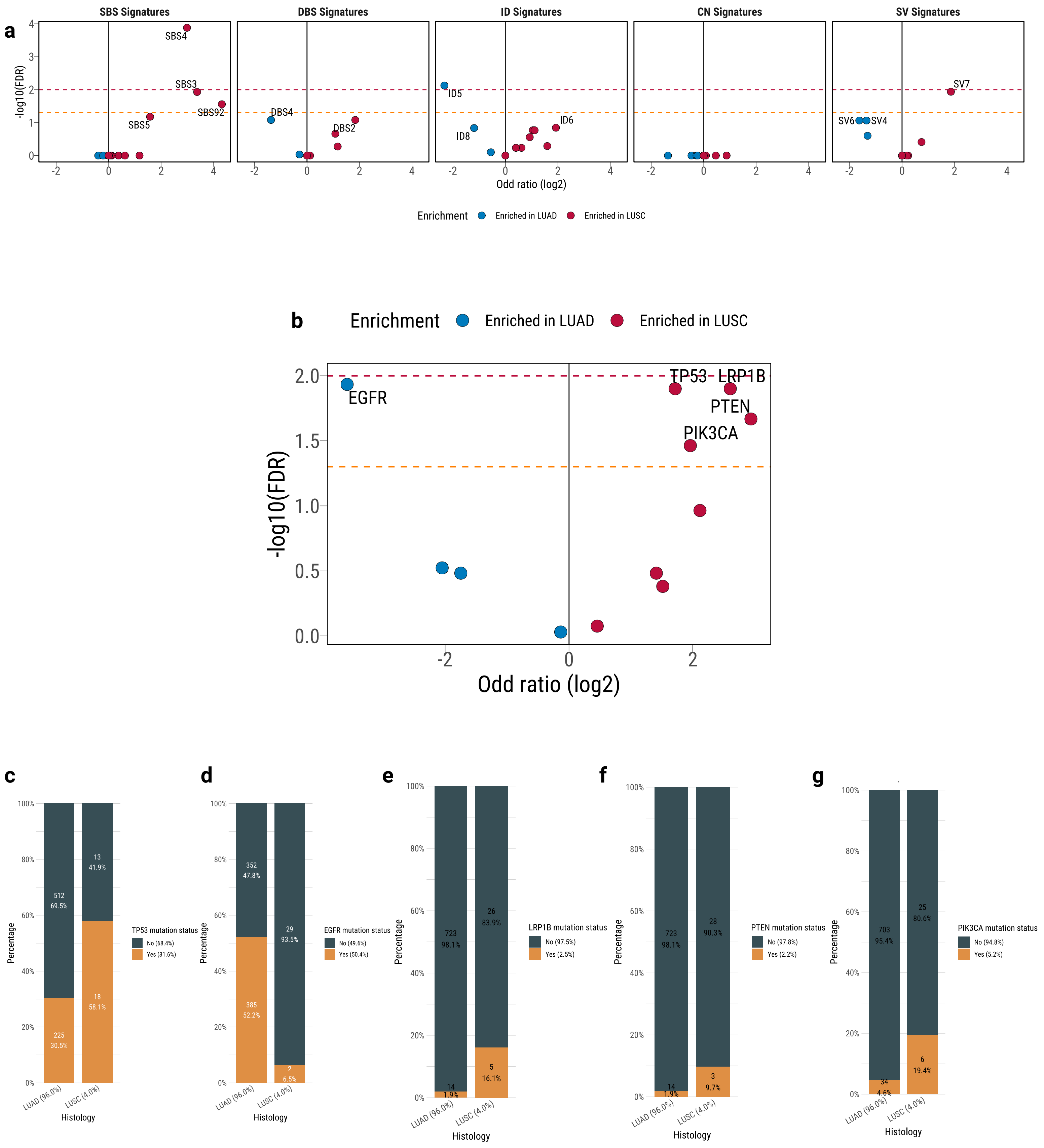

### Extended Data Fig. 7

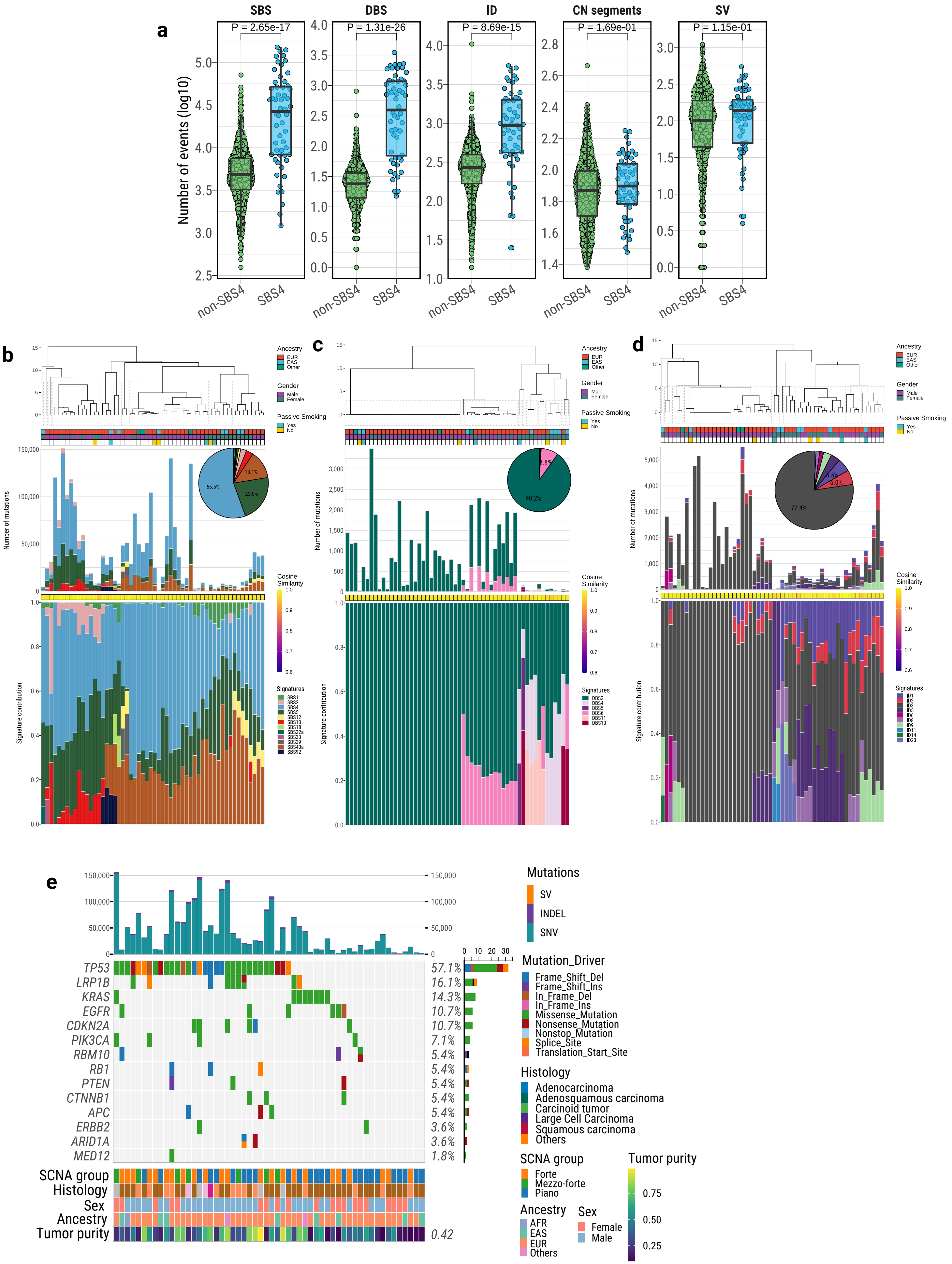

### Extended Data Fig. 8

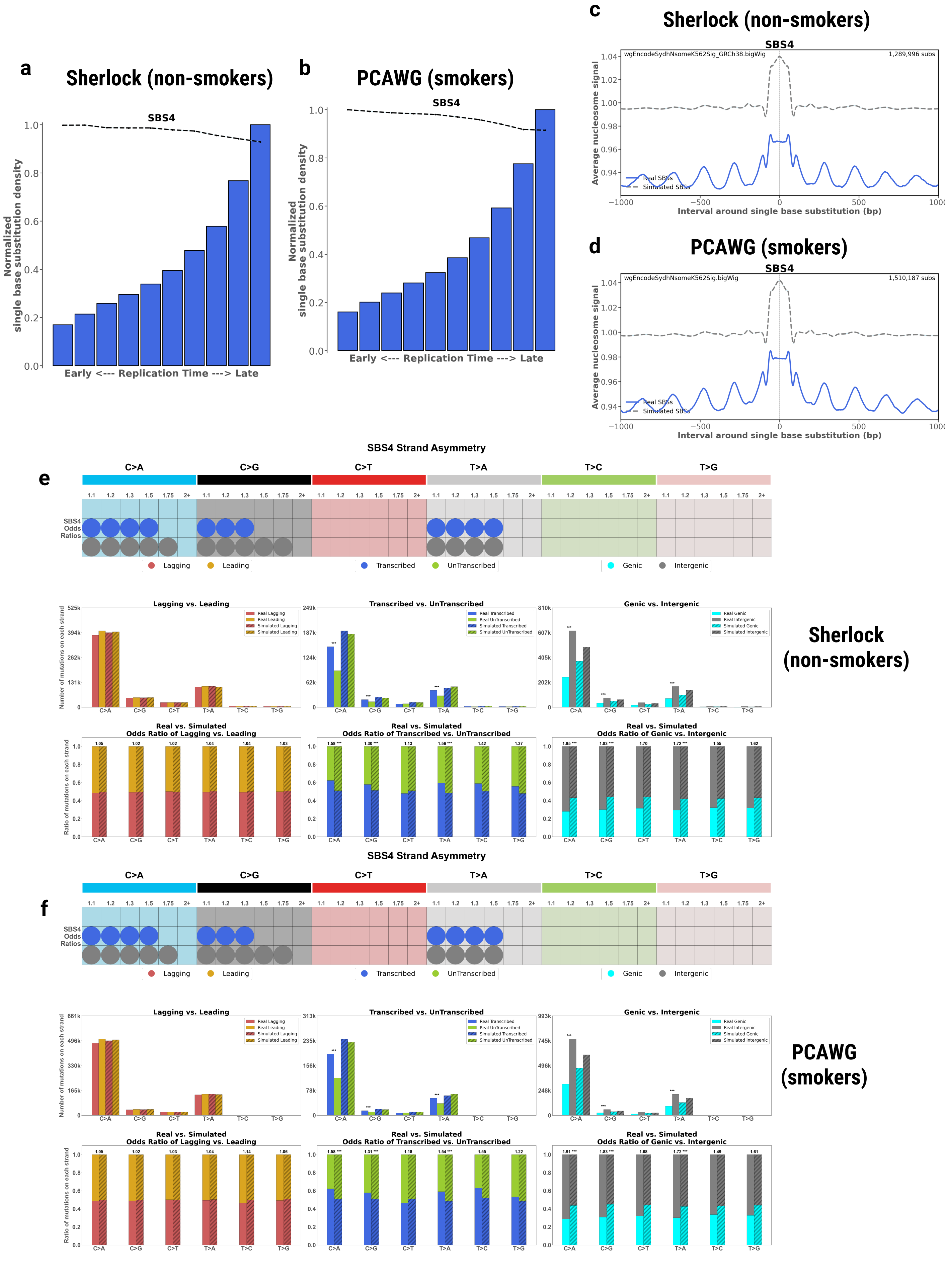

### Extended Data Fig. 9

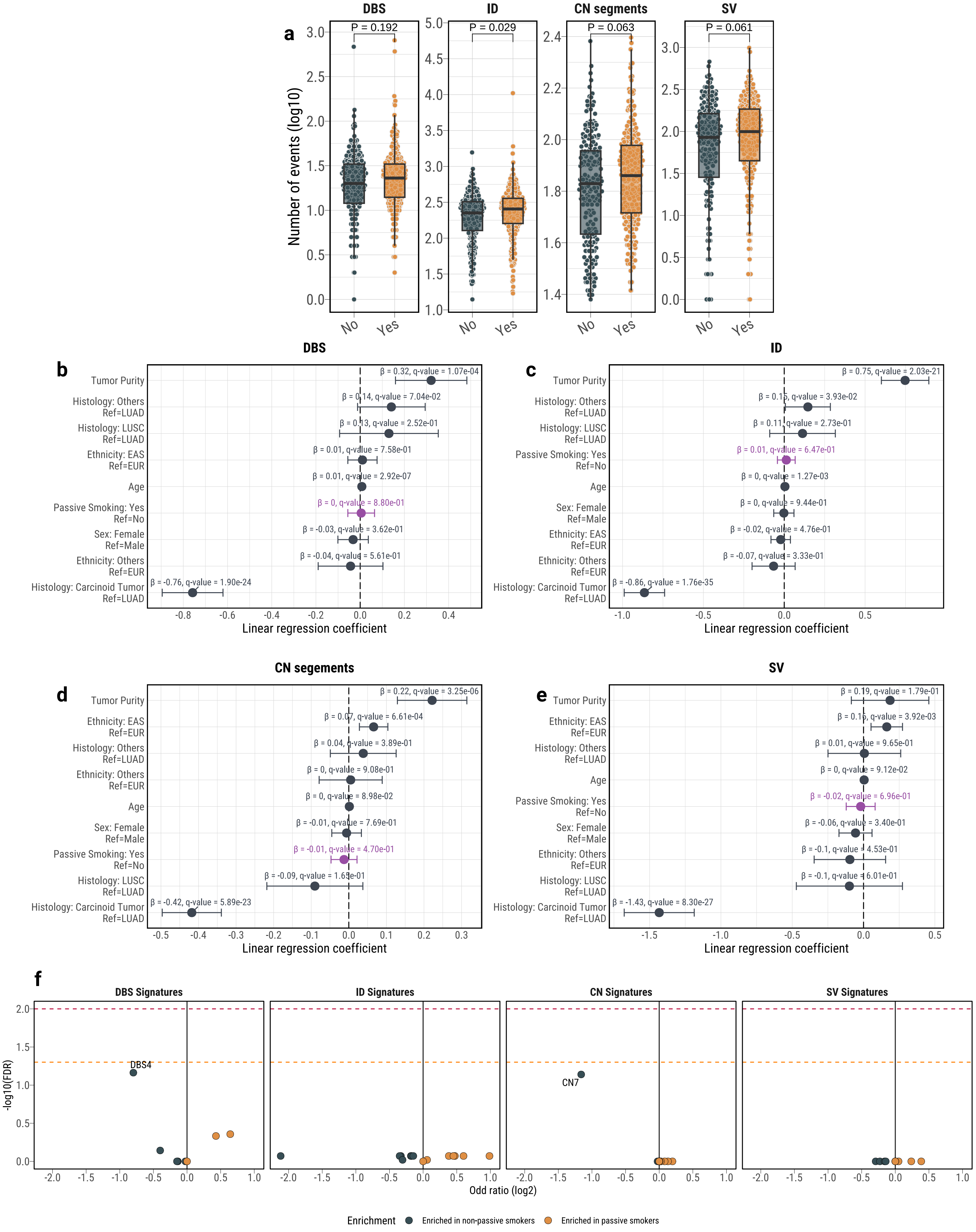

#### Extended Data Fig. 10

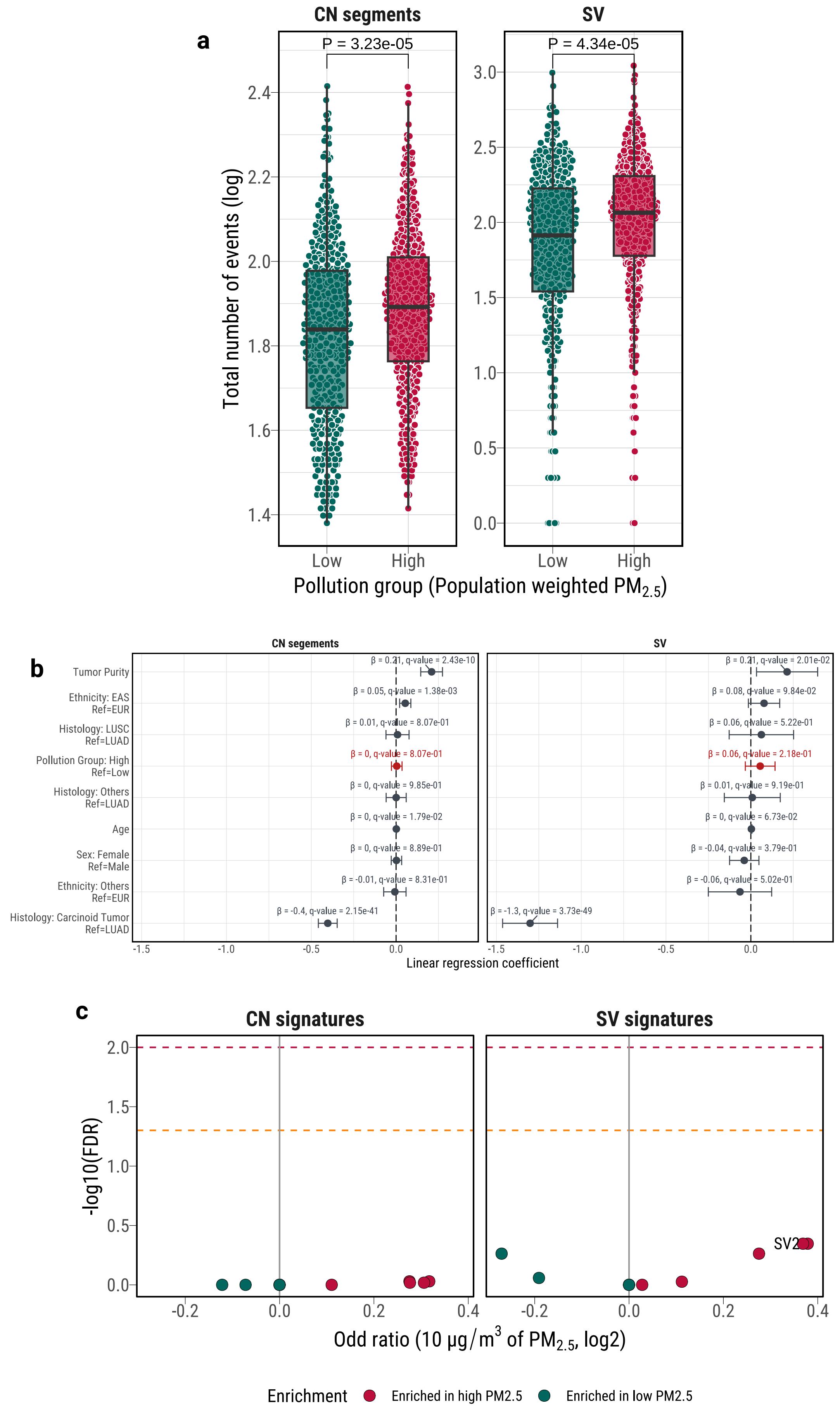

**Extended Data Fig. 11**

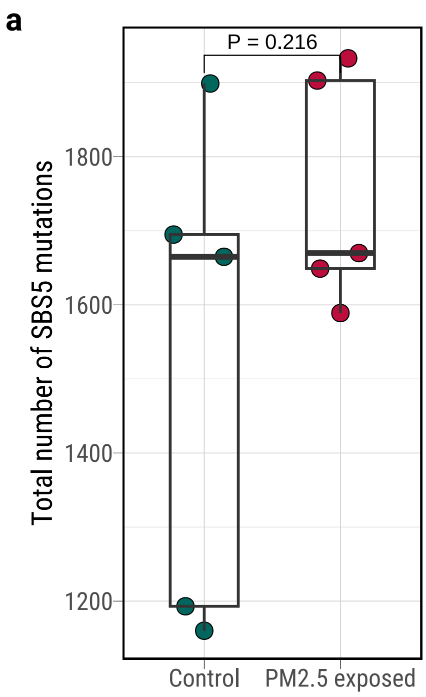

### Extended Data Fig. 12

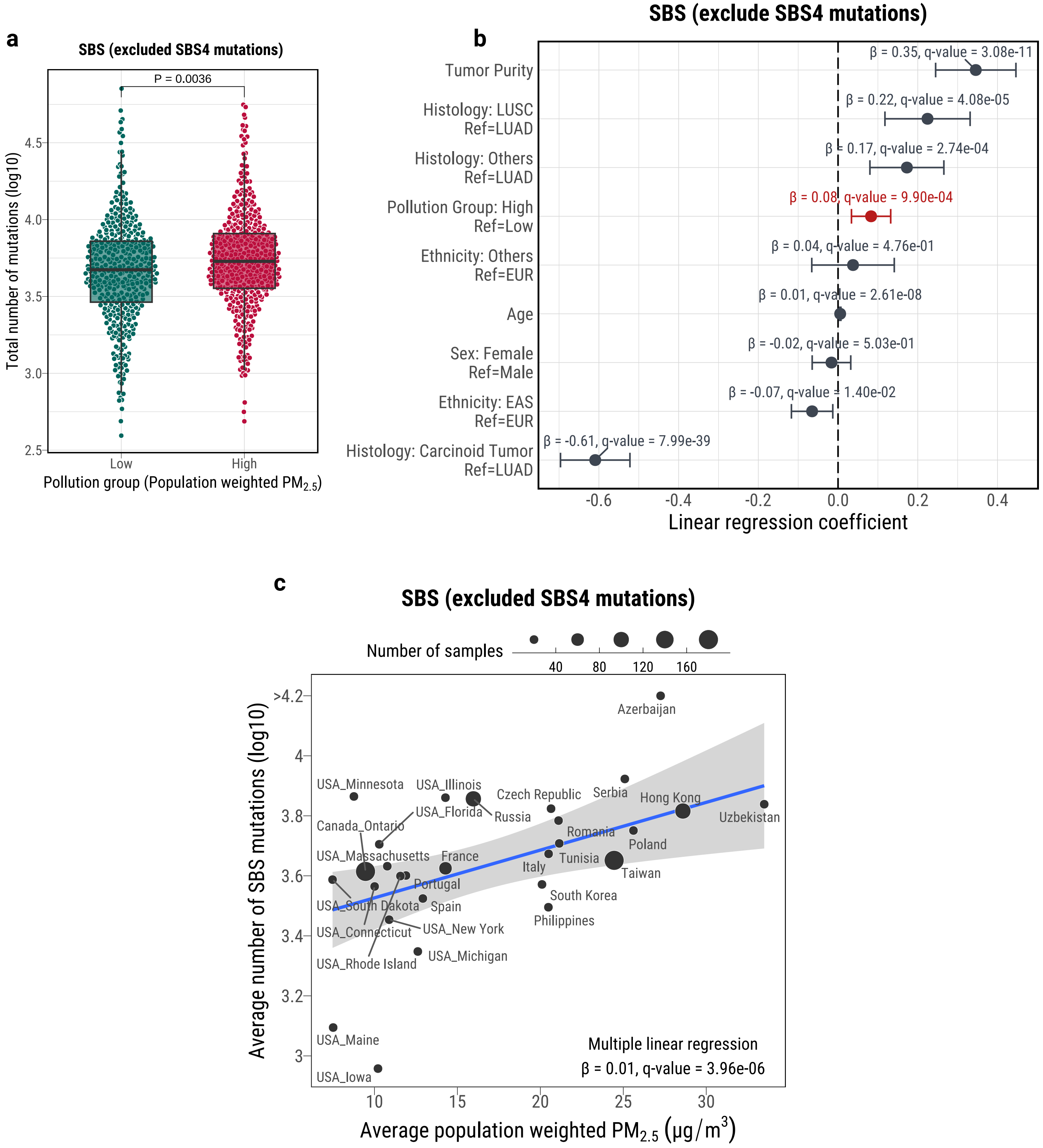
