## Supplementary Figures for "The mutagenic forces shaping the genomic landscape of lung cancer in never smokers"

### Supplementary Fig. 1

a

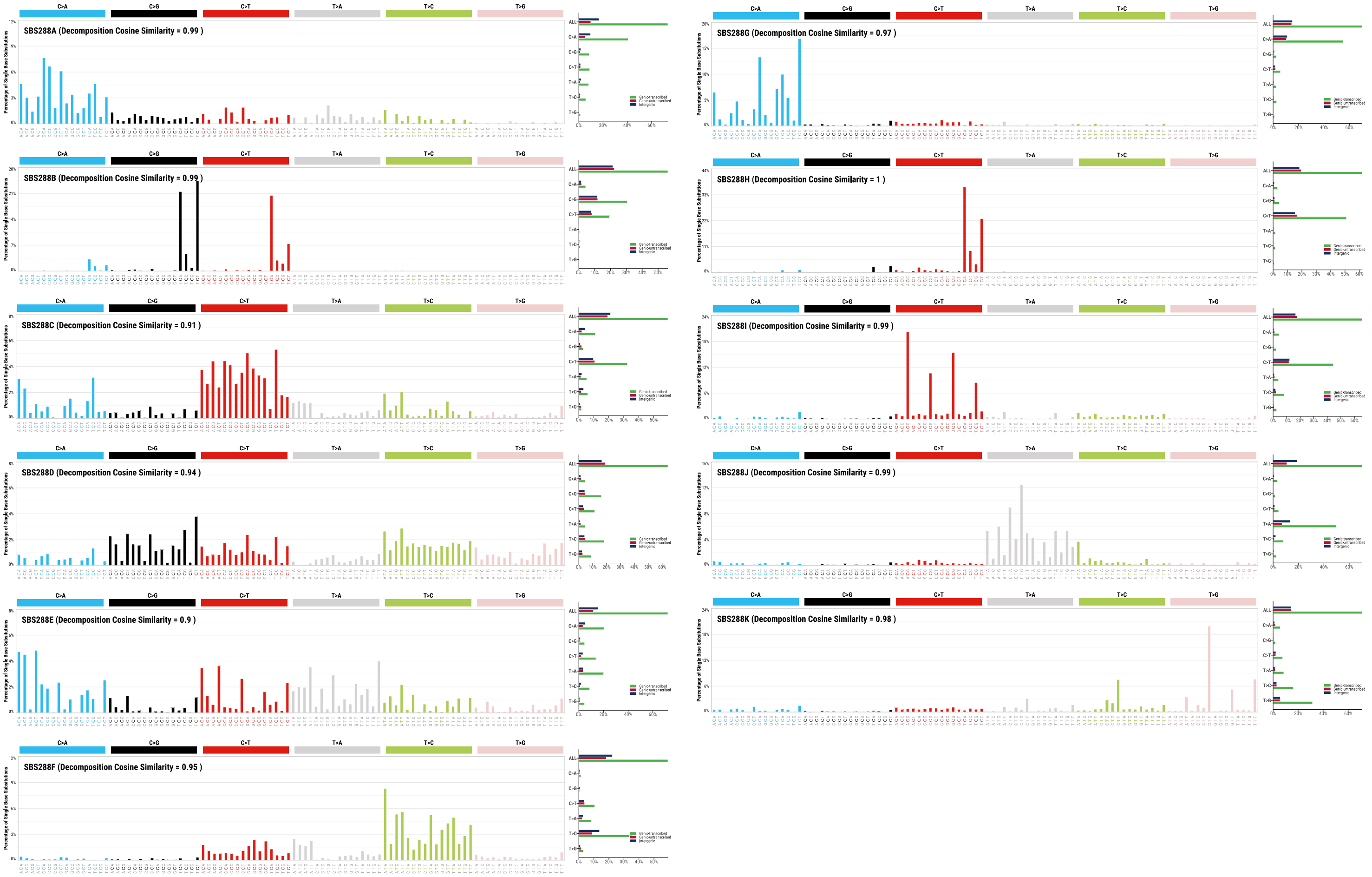


b

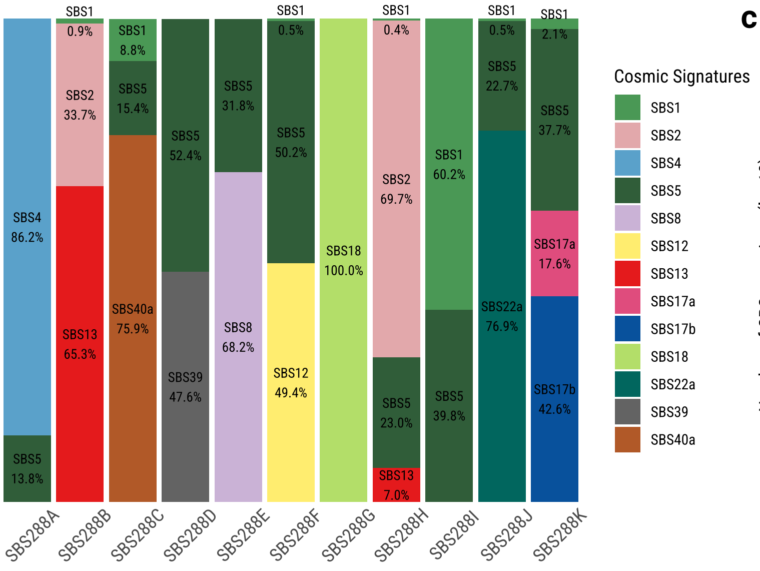


c

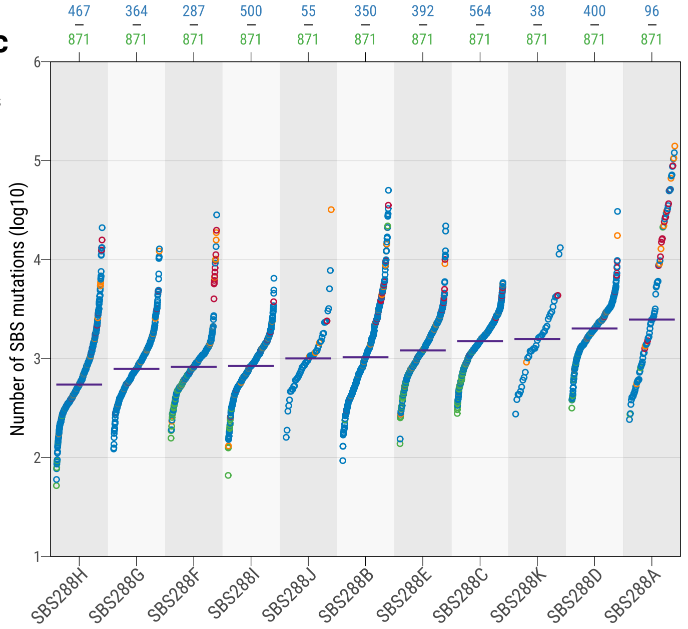

### Supplementary Fig. 2

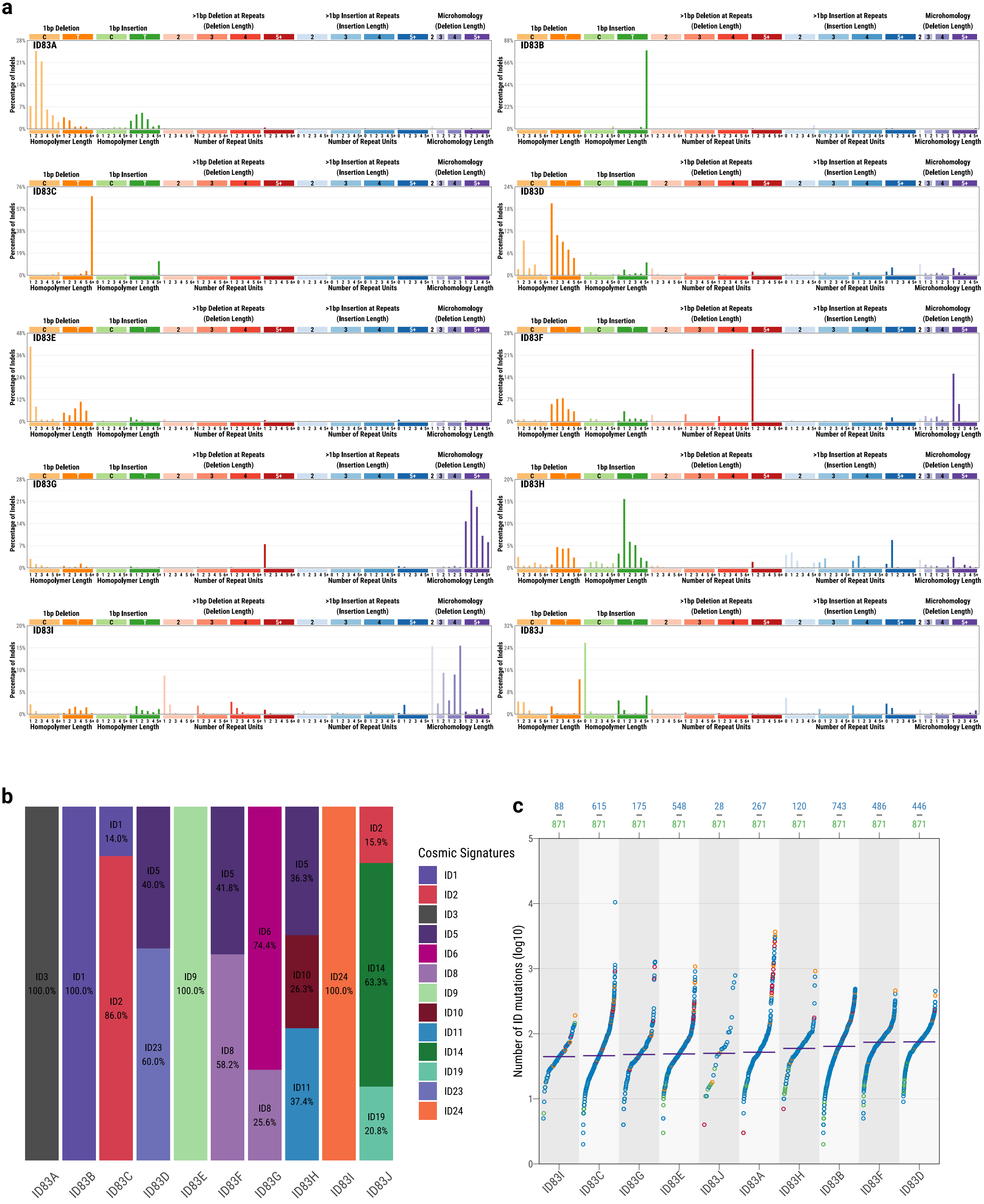

#### Supplementary Fig. 3

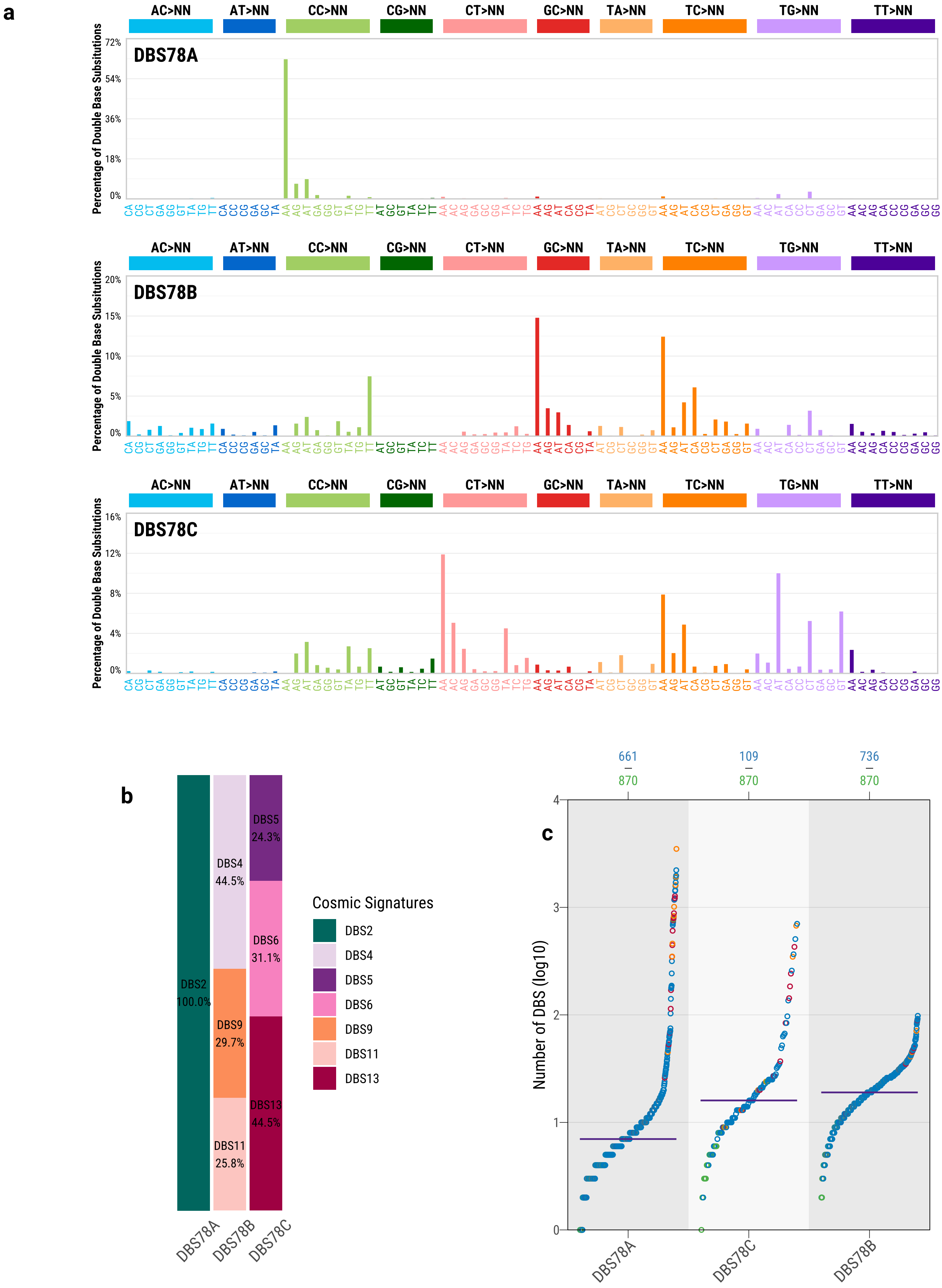

#### Supplementary Fig. 4

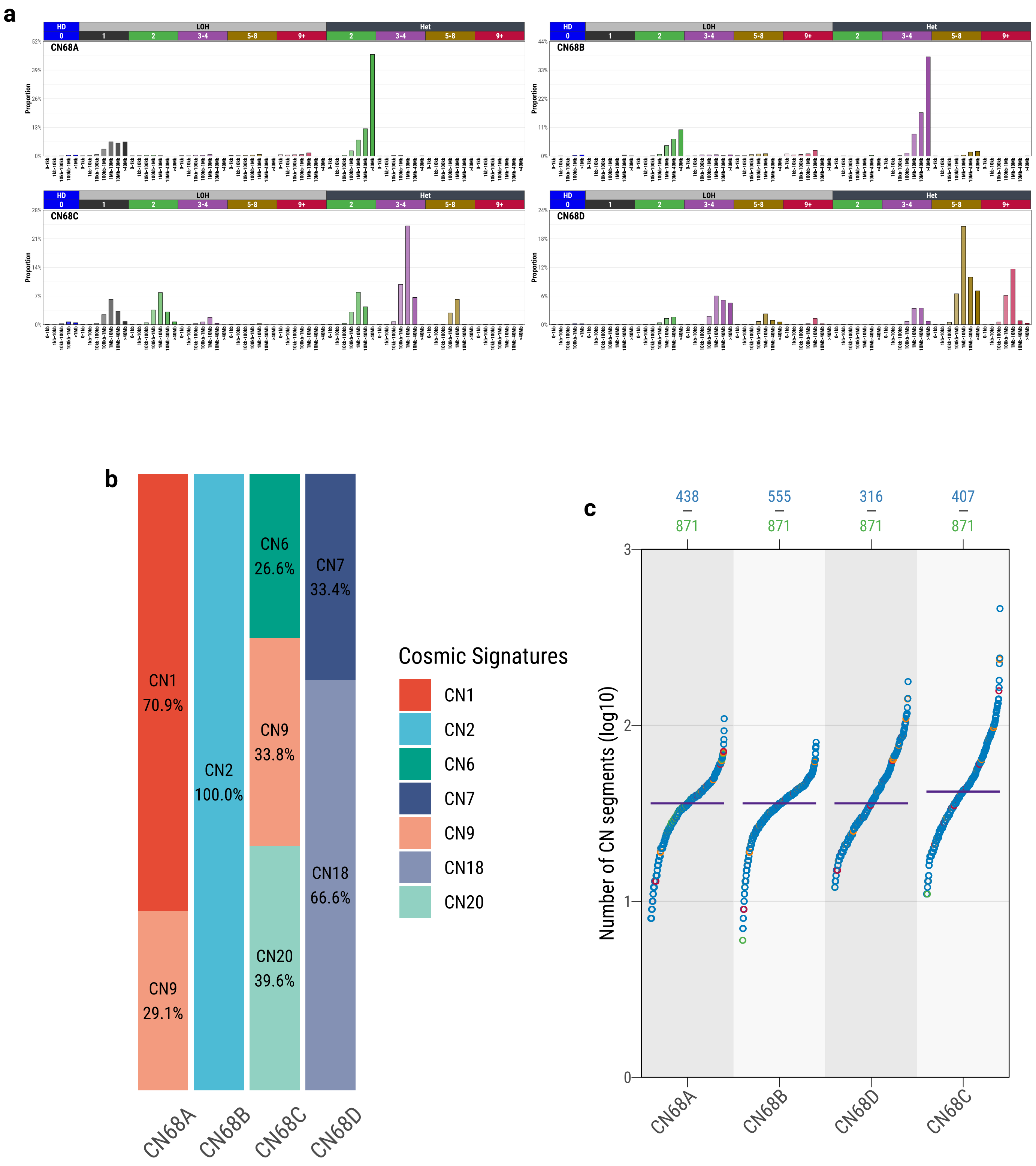

#### Supplementary Fig. 5

**a**

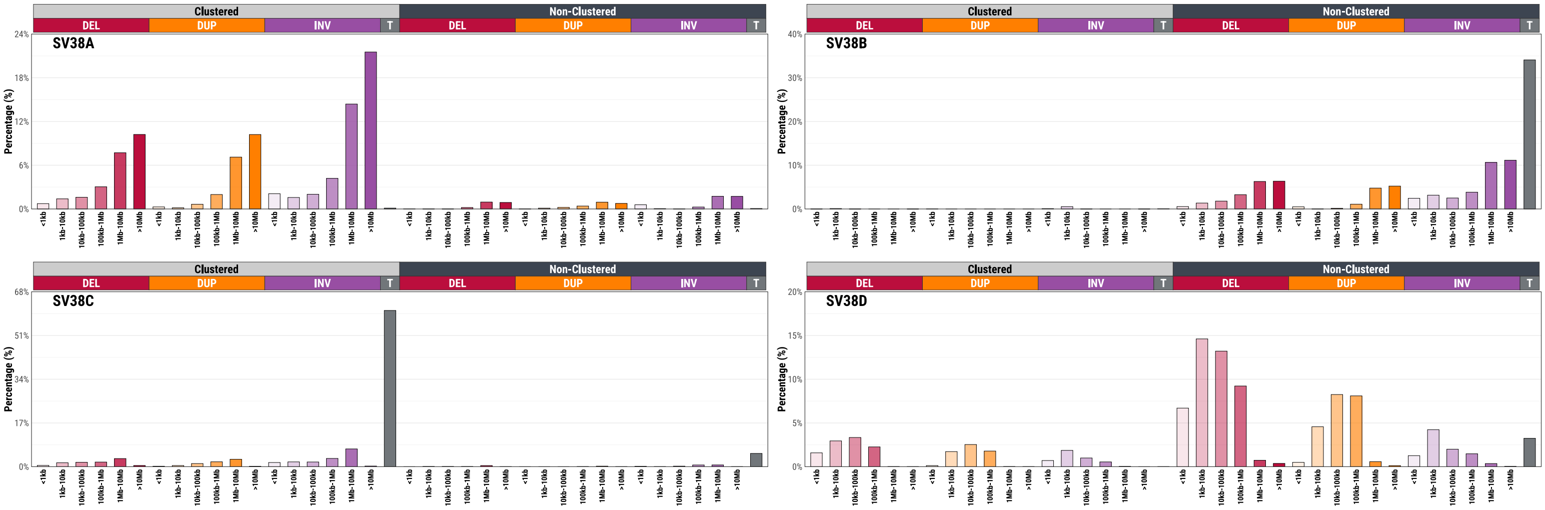


**b**

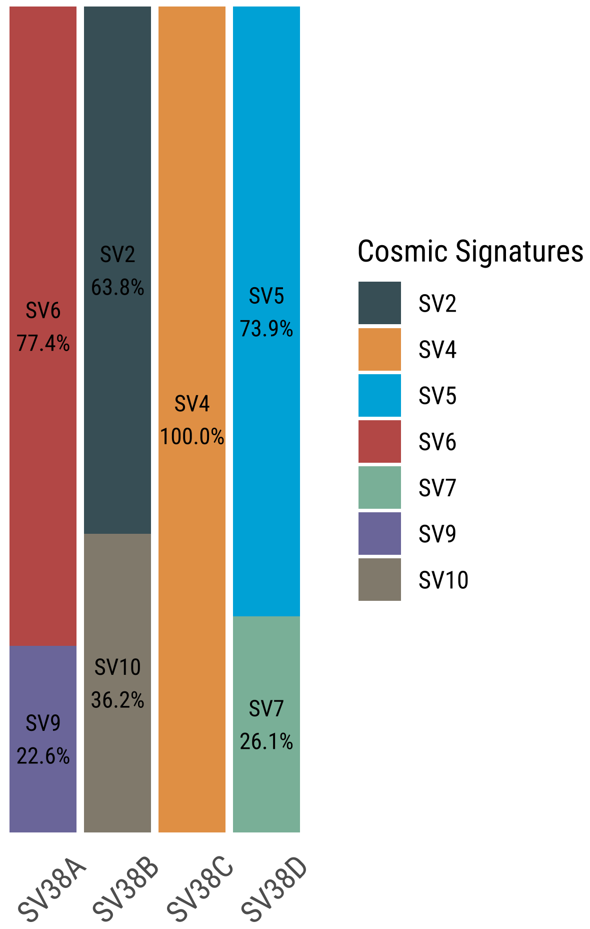


**c**

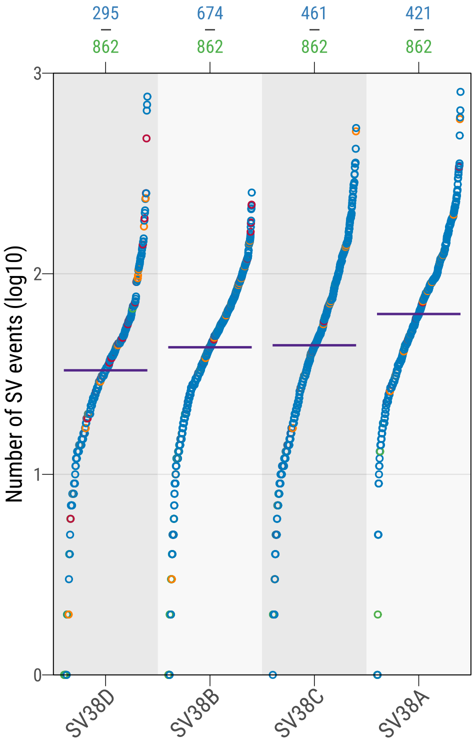

**Supplementary Fig. 6**

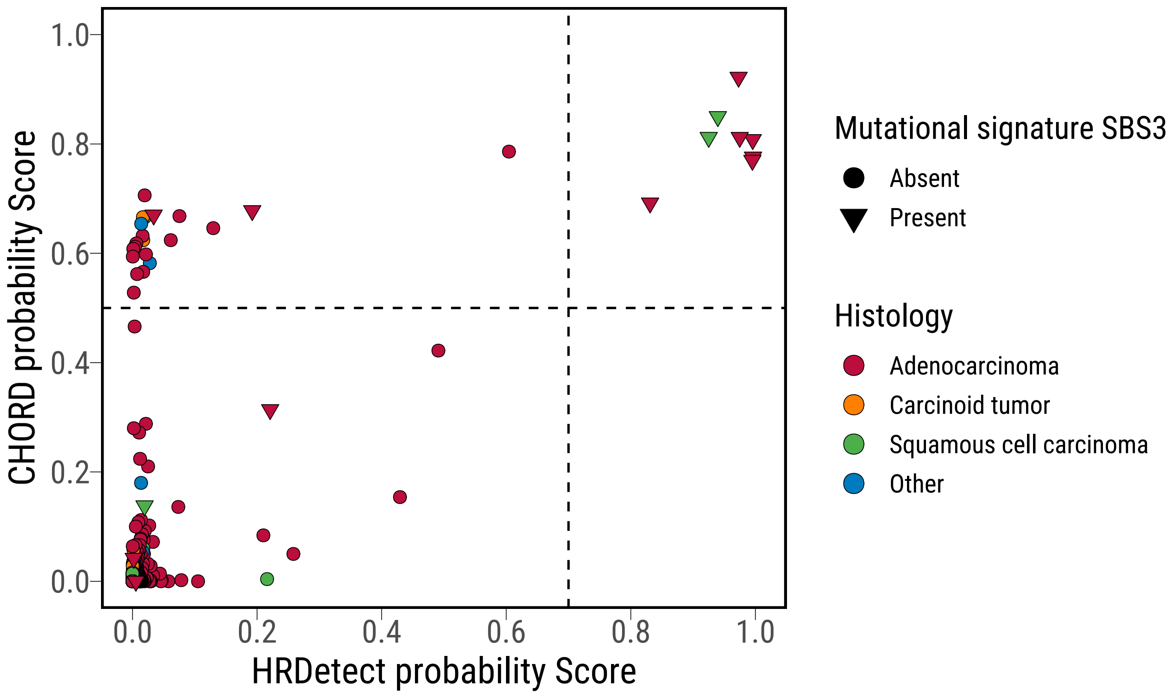

**Supplementary Fig. 7**

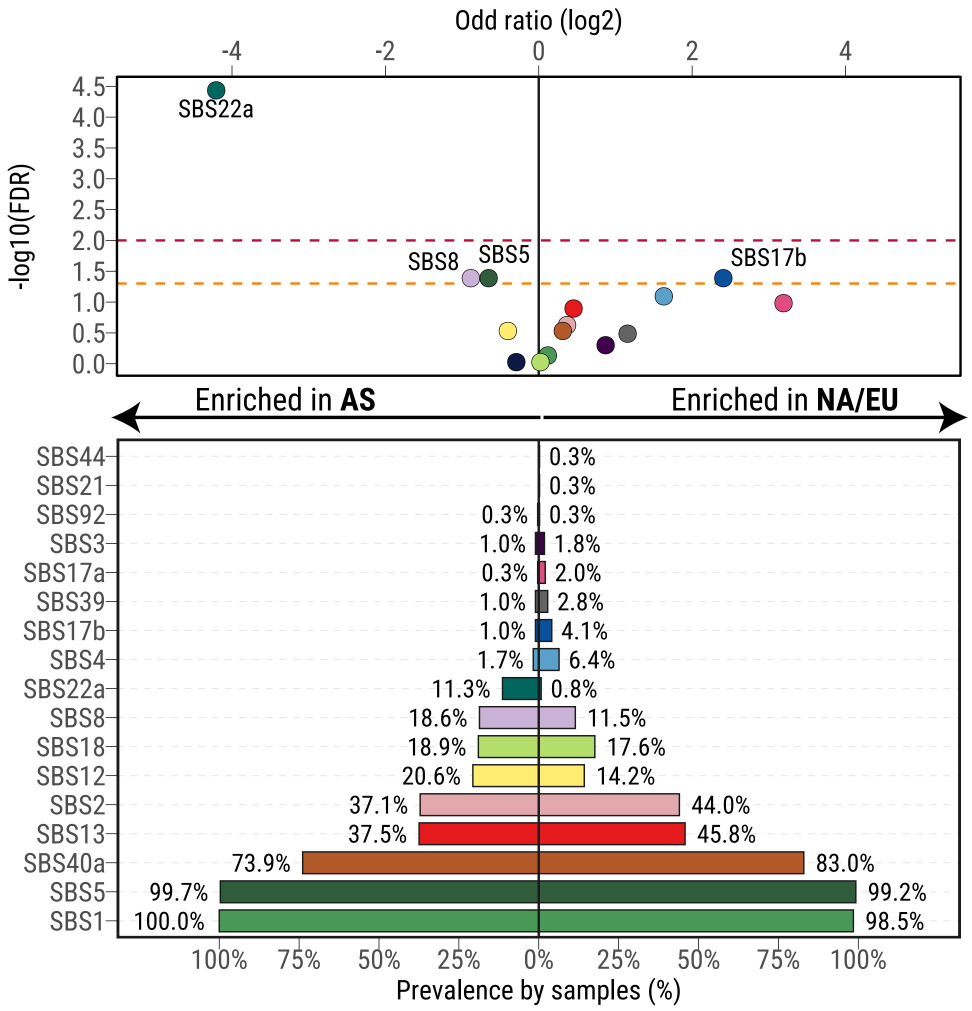

#### Supplementary Fig. 8

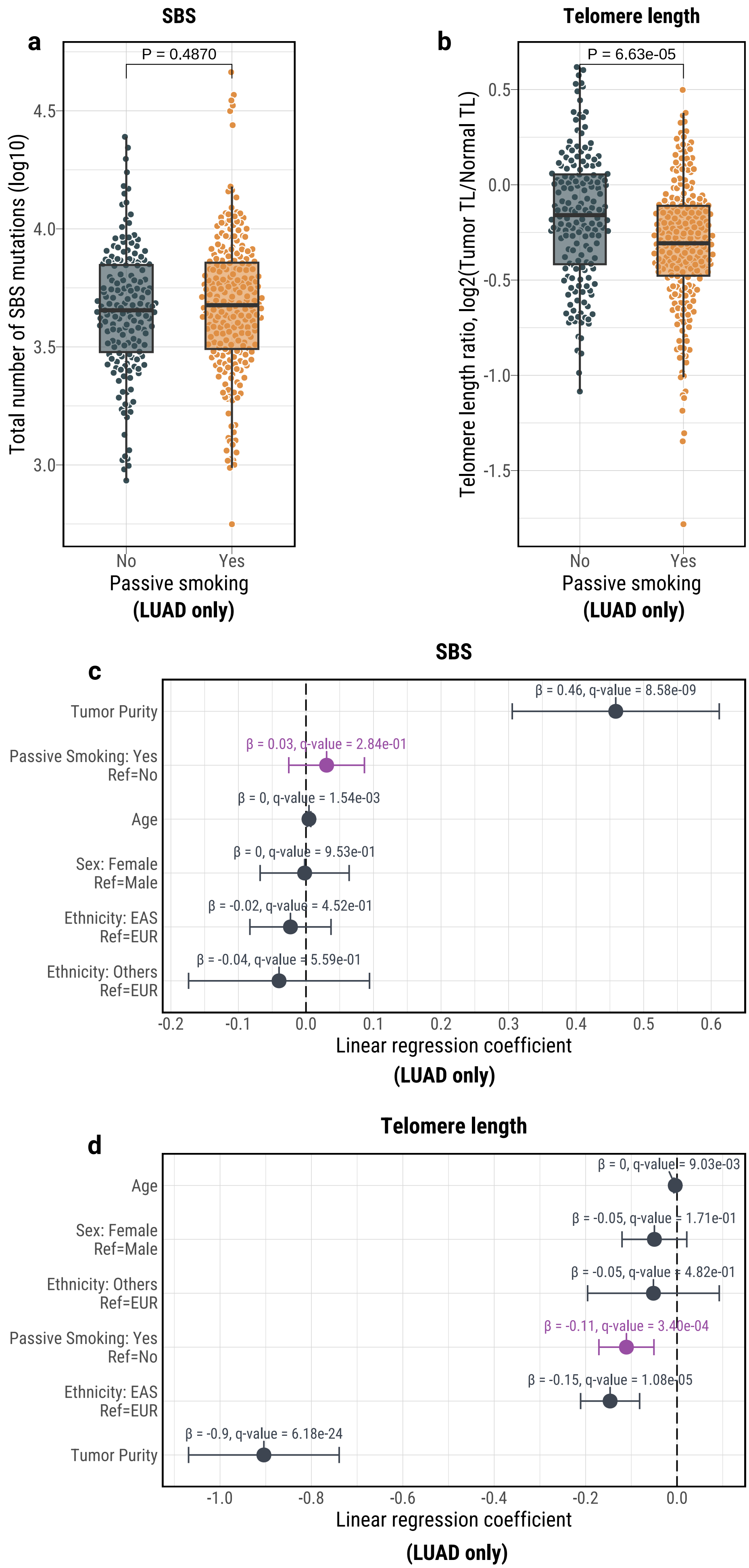

**Supplementary Fig. 9**

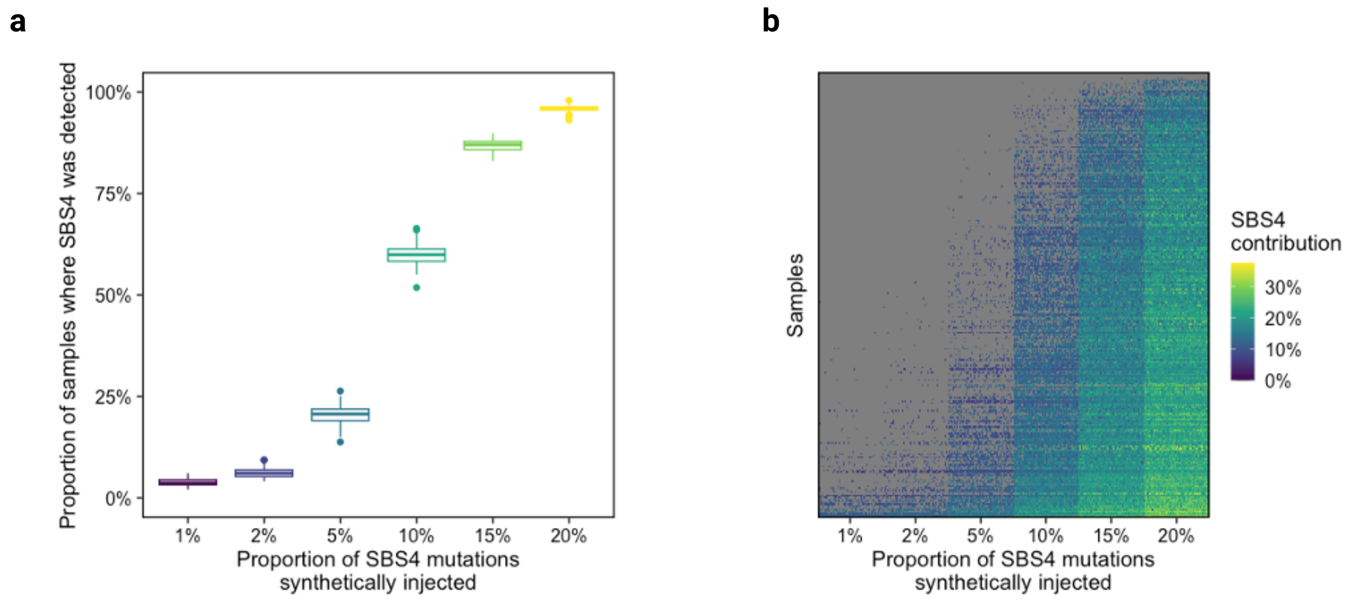

#### Supplementary Fig. 10

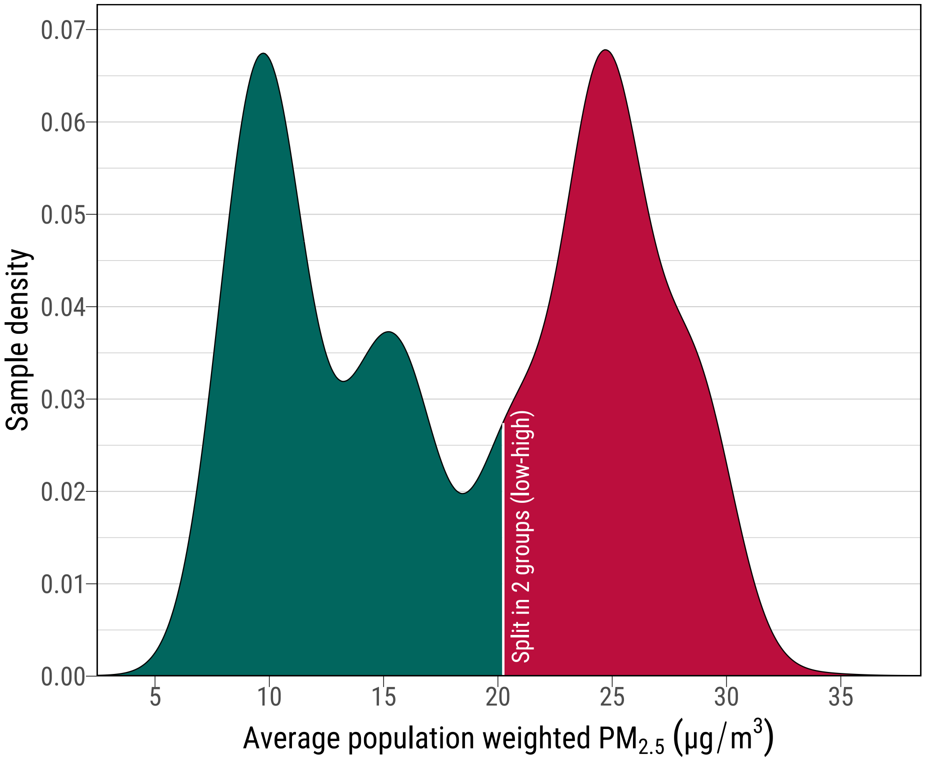
